# Test-retest reliability of intrahemispheric dorsal premotor and primary motor cortex dual-site TMS connectivity measures

**DOI:** 10.1101/2024.03.14.24304288

**Authors:** Robin E. Heemels, Sian Ademi, Melina Hehl

## Abstract

**Objective:** Investigating the optimal interstimulus interval (ISI) and the 24-hour test-retest reliability for intrahemispheric dorsal premotor cortex (PMd) – primary motor cortex (M1) connectivity using dual-site transcranial magnetic stimulation (dsTMS).

**Methods:** In 21 right-handed adults, left intrahemispheric PMd–M1 connectivity has been investigated with a stacked-coil dsTMS setup (conditioning stimulus: 75% of resting motor threshold; test stimulus: eliciting MEPs of 1–1.5 mV) at ISIs of 3, 5–8, and 10 ms. Additionally, M1–M1 short-interval intracortical inhibition (SICI) and intracortical facilitation (ICF) were investigated to assess comparability to standard paired-pulse setups.

**Results:** Conditioning PMd led to significant inhibition of M1 output at ISIs of 3 and 5 ms, whereas 10 ms resulted in facilitation (all, p < 0.001), with a fair test-retest reliability for 3 (ICC: 0.47) and 6 ms (ICC: 0.44) ISIs. Replication of SICI (p < 0.001) and ICF (p = 0.017) was successful, with excellent test-retest reliability for SICI (ICC: 0.81).

**Conclusion:** This dsTMS setup can probe the inhibitory and facilitatory PMd–M1 connections, as well as reliably replicate SICI and ICF paradigms.

**Significance:** The stacked-coil dsTMS setup for investigating intrahemispheric PMd–M1 connectivity offers promising possibilities to better understand motor control.

## 1. INTRODUCTION

During the execution of voluntary movement, the primary motor cortex (M1) as well as the premotor and supplementary motor areas play an important role (Shibasaki, 2012, Welniarz et al., 2019). For example, the planning, execution, and control of voluntary movements require a well-coordinated co-activation of M1 and the dorsal premotor cortex (PMd), which is located 0.8–2.3 cm anteriorly of M1 (Li et al., 2015, Picard and Strick, 2001). More specifically, signals emerging from M1 initiate the movement (Tracy et al., 1998), whereas the premotor cortex, among other regions, generates and updates movement plans through higher-order visuomotor processing, incorporating feedback from M1 and other brain areas (e.g., dorsolateral prefrontal cortex, supplemental motor area (SMA), and parietal cortex) (Abe and Hanakawa, 2009, Beets et al., 2015, Fukushima et al., 2018, Grafton et al., 1998, Halsband and Passingham, 1985, Nakayama et al., 2008, Rushworth et al., 2003). In case one or both of these brain areas are impaired, e.g., due to a stroke, a range of motor deficits such as weakness, spasticity, as well as compromised movement control and coordination can be observed, highlighting the importance of this PMd–M1 connectivity for motor control (Stewart et al., 2016).

One way to investigate PMd–M1 connectivity is dual-site transcranial magnetic stimulation (dsTMS), a tool that allows studying the effective or causal functional connectivity between two brain areas at rest, but also during task preparation or execution within a temporal resolution of milliseconds (Van Malderen et al., 2023). To do so, a conditioning stimulus (CS) over a brain area of interest is followed by a test stimulus (TS) over the M1 representation a few milliseconds later, and the peak-to-peak amplitude of these conditioned motor-evoked potentials (MEP) is compared to unconditioned MEPs emerging from M1 (Kujirai et al., 1993). A smaller or larger conditioned MEP as compared to unconditioned MEP, points towards an inhibitory or facilitatory influence of the conditioned brain area on M1, respectively (Van Hoornweder et al., 2021, Van Malderen et al., 2023). However, the nature of this effect is dependent on the stimulation protocol, depending on parameters such as the choice of the CS and TS intensity, the coil placement, and the interstimulus interval (ISI).

While interhemispheric PMd–M1 connectivity at rest has been extensively investigated, and yielded inhibition or facilitation of contralateral M1 output mainly depending on the CS intensity applied over PMd (i.e., suprathreshold or subthreshold respectively) (Bäumer et al., 2009, Koch et al., 2006, Ni et al., 2009, Rothwell, 2011), studying the intrahemispheric PMd–M1 connectivity is challenging due to the proximity of PMd and M1 (Amiez et al., 2006, Fiori et al., 2016, Groppa et al., 2012, Parmigiani et al., 2015, Van Malderen et al., 2023, Vesia et al., 2017). As a result, dsTMS studies that investigated the intrahemispheric PMd–M1 interaction implemented various coil placements and yielded diverse results [for a detailed review, see Van Malderen et al. (2023)]. In summary, prior research regarding the left intrahemispheric PMd–M1 interaction has shown a significant decrease in

MEP amplitude at an ISI of 4 and 6 ms using a CS intensity at 90% of the active motor threshold (aMT). Interestingly, increasing the CS intensity to 120% of the aMT led to facilitation at an ISI of 6 ms (Civardi et al., 2001). However, these findings could not be replicated by Bäumer et al. (2009) who instead showed inhibition of M1 output using a CS intensity at 90% of the aMT at a longer ISI of 8 ms, or with a CS at 110% of the resting motor threshold (rMT) combined with shorter ISIs of 2 and 4 ms. Moreover, highly variable outcomes were observed for protocols with the TS preceding the CS by 2.4–4.4 ms, resulting in facilitation (Groppa et al., 2012). However, when applying the same stimulation parameters but using a different coil setup, these results could not be replicated (Van Hoornweder et al., 2021). Nevertheless, it is worth noting that overall, differences in stimulation protocols, as mentioned previously, complicate comparison between studies. More specifically, in the aforementioned studies (Bäumer et al., 2009, Civardi et al., 2001, Van Hoornweder et al., 2021), interstimulus distances of 3–6 cm have been used, exceeding the average anatomical PMd–M1 distance of 0.8–2.3 mm (Picard and Strick, 2001). This is a direct result of the geometry of standard TMS coils, which are too large to allow for smaller interstimulus distances. In addition, using smaller TMS coils allowing for reduced interstimulus distances can result in significant heating, especially during longer TMS protocols, and occasionally fail to elicit MEPs in participants with higher motor thresholds. As a result, available research investigating intrahemispheric PMd–M1 interactions is characterized by diverse coil placements, and CS locations. Finally, only a small number of ISIs were tested, with varying CS and TS stimulation intensities.

One factor that could affect the variability in the findings mentioned above is the test-retest reliability. This refers to the variation in outcomes caused by measurement errors or short-term changes in participants’ physiology. Prior research concerning the test-retest reliability of commonly employed single-coil paired-pulse TMS (ppTMS) protocols such as short-interval intracortical inhibition (SICI) and intracortical facilitation (ICF) yielded intra-class correlation coefficients (ICC) ranging from 0.23–0.91 for SICI (Biabani et al., 2018, Du et al., 2014, Fleming et al., 2012, Hermsen et al., 2016, Matamala et al., 2018, Ngomo et al., 2012), whereas for ICF lower ICCs of 0.04–0.82 were reported (Biabani et al., 2018, Du et al., 2014, Hermsen et al., 2016). In contrast, dsTMS protocols showed even lower test-retest reliability with ICCs, ranging from 0.01–0.69 for the SMA–M1 interaction (Rurak et al., 2021, 2022). So far, there is limited research on the reliability of PMd–M1 dsTMS measures although sufficient test-retest reliability is essential for future dsTMS studies. As a solution to the diverse coil placements with large interstimulus distances, we recently introduced and validated a novel dsTMS setup using two stacked figure-of-eight TMS coils (Hehl et al., 2024). This setup allows us to investigate the intrahemispheric PMd–M1 interaction with an interstimulus distance of 2.1 cm while using off-the-shelf TMS equipment.

Using the aforementioned dsTMS setup with the optimized coil placement to target PMd and M1, the objectives of the present study were threefold. First, we investigated the intrahemispheric PMd–M1 interaction with a stacked dsTMS setup, and more specifically the effects of different ISIs on M1 output. Second, we measured the test-retest reliability of PMd–M1 interaction measurements for each of these ISIs. Lastly, to allow for a comparison of the obtained PMd–M1 reliability results with well-established single-coil ppTMS protocols, additional SICI and ICF measurements were obtained with a slightly adapted dsTMS setup (i.e., aligning the coils’ focal stimulation points).

## 2. METHODS

### 2.1. Participants

Twenty-one participants (10 female, 11 male) aged 22.5 ± 2.6 years [mean ± standard deviation (SD)] participated in the study (see Table 1). All participants were screened for right-handedness [Edinburg Handedness Inventory (EHI): 80.0 ± 18.4, Oldfield (1971)], normal cognition [Montreal cognitive assessment (MoCA): 29.0 ± 1.1, Nasreddine et al. (2005)], and no self-reported symptoms of depression [Beck Depression Inventory (BDI–II): 4.4 ± 3.8, Beck et al. (1996)]. Moreover, participants were screened for the absence of central nervous system or psychiatric disorders, neuroactive medication use, history of brain injury or surgery, and contraindications for TMS (Wassermann, 1998). All participants provided written informed consent and completed the study protocol without the occurrence of adverse effects. This study was approved by the Ethics Committee Research UZ/KU Leuven (S65077) and in compliance with the declaration of Helsinki (World Medical Association, 2013).

**Table 1.** Study sample characteristics.

| n = 21 (11M, 10F) | Min | Max | Median | Mean | SD |
| --- | --- | --- | --- | --- | --- |
| Age (years) | 19.0 | 31.0 | 22.0 | 22.5 | 2.6 |
| EHI LQ (%) | 50.0 | 100.0 | 80.0 | 80.0 | 18.4 |
| MoCA | 27.0 | 30.0 | 29.0 | 29.0 | 1.1 |
| BDI-II | 0.0 | 12.0 | 3.0 | 4.4 | 3.8 |
| Head circumference (cm) | 53.5 | 59.5 | 57.0 | 56.8 | 1.4 |
| Nasion inion distance (cm) | 32.8 | 39.5 | 36.0 | 36.2 | 1.6 |
| Pre-auricular distance (cm) | 32.8 | 37.5 | 36.0 | 35.3 | 1.5 |

*Abbreviations:* BDI-II = Beck Depression Inventory; EHI LQ = laterality quotient assessed by Edinburg handedness inventory; MoCA = Montreal cognitive assessment; M = male; F = female; SD = standard deviation.

### 2.2. Electromyography preprocessing and stereotactic navigation

Electromyographic (EMG) activity was recorded from the cleaned and gently abraded (3M™ Red Dot™ Trace Prep 2236, 3M Health Care, St. Paul, MN, USA) muscle belly of both right and left first dorsal interosseus (FDI) using surface Ag-electrodes (Bagnoli DE-2.1 EMG sensors, DELSYS Inc, Boston, MA, USA). Reference electrodes were placed bilaterally on the ulnar bone of the dorsal wrist. Raw EMG signals were collected (Bagnoli-4 EMG System, DELSYS Inc), filtered for 50/60 Hz noise (HumBug, Quest Scientific, North Vancouver, BC, Canada), amplified (gain = 1000), bandpass filtered (20–2000 Hz), and digitized at 5000 Hz (CED 1401 micro, CED Limited, Cambridge, UK) before being stored on a computer for offline processing. Throughout the experiment, participants were asked to keep both hands relaxed and their eyes open.

### 2.3. Transcranial magnetic stimulation

Dual-site and paired-pulse biphasic TMS pulses were delivered through MC-B70 and MC-B65HO figure-of-eight coils (MagVenture A/S, Farum, Denmark, MC-B70: 150° angled, outer coil winding diameter 97 mm; MC-B65HO outer coil winding diameter 75 mm) connected to MagPro X100 stimulators (MagVenture A/S, Farum, Denmark). Accurate coil placement, i.e., 45° rotated away from the midsagittal line and tangential to the scalp inducing a posterior-to-anterior-directed current, was confirmed using Brainsight®2 neuronavigation software (Rogue Research Inc, Montreal, QC, Canada) and a Polaris Vicra optical tracker (Northern Digital Inc., Waterloo, ON, Canada), based on the Montreal Neurological Institute (MNI) average head brain model. A standardized procedure was used to determine the hotspot of the left M1 (Hehl et al., 2020). A virtual 19×19, 1 cm-spaced grid was centered around the vertex [determined according to the EEG 10-20 system, Klem et al. (1999)] and aligned with the virtual scalp surface using Brainsight®2. The hotspot was defined as the location that elicited the strongest and most consistent MEPs, averaged over five consecutive pulses with the MC-B70 coil. Next, the resting motor threshold (rMT) was defined as the lowest stimulus intensity eliciting at least five out of ten MEPs ≥ 50 µV peak-to-peak amplitude. The rMT was determined for both the MC-B70 and MC-B65HO coil. Lastly, the TS intensity was determined as the lowest intensity eliciting stable MEPs of 1–1.5 mV, averaged over five consecutive pulses using the MC-B70 coil. During these preparatory measurements, trials exceeding a root mean square (RMS) of 5 µV in the 100–50 ms preceding the TMS pulses were discarded, and measurements were repeated.

### 2.4. Dual-site and paired-pulse TMS

The intrahemispheric modulation of M1 by a preceding CS was probed using a stacked dsTMS set-up using two overlapping figure-of-eight coils, MC-B65HO and MC-B70 to apply CS and TS respectively. The MC-B65HO coil is capable of transmitting pulses through the 1.55 cm thin middle section of the angled MC-B70 coil [see Figure 1A and 1B; more detailed visualization in Hehl et al. (2024)]. This enables the stimulation of the PMd and M1 with an interstimulus distance of 2.1 cm, further referred to as dsTMS (see Figure 1A). Additionally, the two coils’ focal stimulation points were aligned to mimic the classic ppTMS setups (see Figure 1B). The setup is described in detail in Hehl et al. (2024). For both dsTMS (PMd–M1) and ppTMS (M1–M1), the TS over M1 was delivered using the MC-B70 coil at an intensity eliciting stable MEPs of 1–1.5 mV averaged over 5 consecutive pulses. The CS intensity was determined based on the rMT of the MC-B65HO coil and upscaled by a correction factor to account for the increased coil-to-cortex distance introduced by stacking the coils: rMT_corrected_ = rMT_MC-B65HO_ x 2.1 [please consult Appendix 1 in Hehl et al. (2024) for details regarding this calculationHehl et al. (2024)].

**Figure 1.**
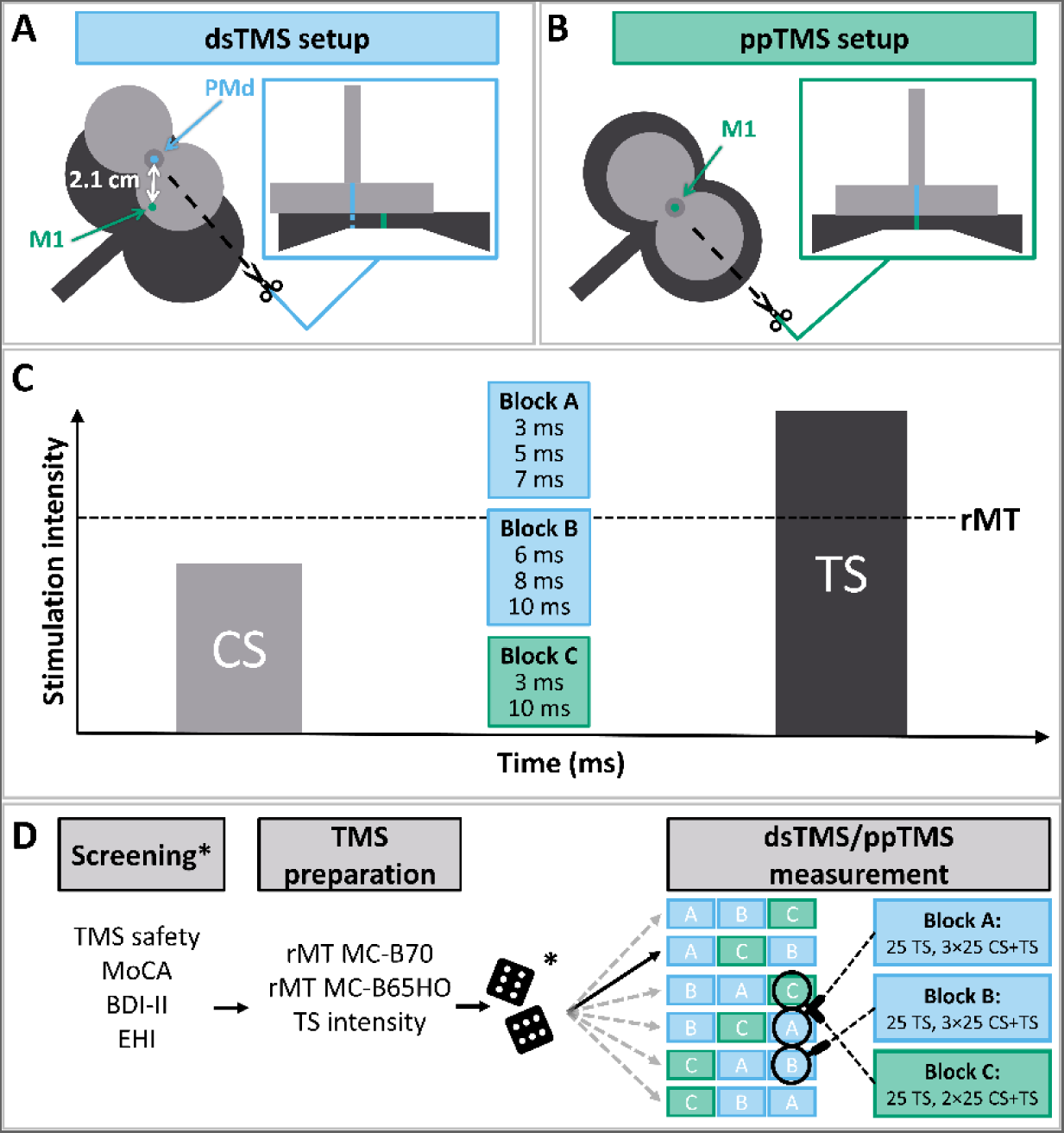
Experimental design and setup**. [A]** The MC-B65HO coil delivering the conditioning stimulus (CS, light gray) placed on top of the MC-B70 coil delivering the test stimulus (TS, dark gray) with an interstimulus distance of 2.1 cm for probing PMd–M1 connectivity. **[B]** The MC-B65HO coil delivering the CS (light gray) placed on top of the MC-B70 coil delivering the TS (dark gray) with an interstimulus distance of 0 cm for probing SICI and ICF. The cross-sectional views of the two coils in **[A]** and **[B]** respectively indicate the most focal stimulation point for each coil. **[C]** Organization of interstimulus intervals (ISI) into three pseudo-randomly presented experimental blocks. Stimulus intensity of CS at 75% of the resting motor threshold (rMT) and TS at an intensity to elicit motor evoked potentials of 1–1.5 mV. **[D]** Flowchart of standardized experimental protocol. Screening participants, determining stimulation intensities, randomizing block order, and measuring dsTMS and ppTMS. Asterisks indicate steps performed during session 1 only.

### 2.5. Experimental protocol

Participants were seated facing a computer screen with their hands and pronated forearms supported by a table. During the measurement blocks, participants were blinded from EMG signals and viewed a landscape slideshow (30 seconds per image) to promote stable excitability.

To determine the optimal ISI for assessing the intracortical connectivity between PMd and M1, two experimental blocks were conducted, each with a duration of ±10 min. Each block consisted of 25 TS trials (stimulating M1), and 25 CS+TS trials (stimulating respectively PMd and M1) for three different ISIs per block (i.e., block A: 6, 8, and 10 ms; block B: 3, 5, and 7 ms), resulting in a total of 100 pseudo-randomized trials (see Figure 1C and 1D). In addition, SICI and ICF were assessed to evaluate the effectiveness and reliability of this dsTMS setup and allow for comparison with earlier reliability work. To do so, a third block with a duration of ±7 min was assessed (block C), consisting of 25 TS trials, and 25 CS+TS trials (all stimulating M1) for an ISI of 3 ms (SICI) and 10 ms (ICF), respectively (75 pseudo-randomized trials in total). The inter-trial interval for all blocks (A, B, and C) was set to 6 s ± 10%. The order of the three experimental blocks (block A and B for dsTMS, block C for ppTMS) was randomized for each participant (see Figure 1D).

### 2.6. Test-retest reliability

To assess the 24-hour test-retest reliability of dsTMS and ppTMS measurements, participants completed the protocol twice. Measurements were scheduled exactly 24 hours apart to minimize circadian rhythm effects on the cortical excitability (Ly et al., 2016). To further minimize the introduced variability between the two measurement sessions, within each participant the (1) Brainsight®2 coil calibrations, (2) randomized experimental block order, (3) the timing of the breaks, and (4) the researcher conducting the measurements were kept constant. Furthermore, participants were asked to maintain and document their usual physical activity levels, sleeping behavior, and consumption of caffeine and alcohol between measurement days.

### 2.7. Data processing

Processing of right and left FDI EMG data was performed in MATLAB® (R2020a, The MathWorks Inc, Natick, MA, USA). Trials containing an EMG RMS ≥ 20 µV during the 100–50 ms preceding the stimulation were removed from the analysis (Cuypers et al., 2020, Hehl et al., 2020). The ratios of the mean dual-site and paired-pulse TMS amplitudes as compared to the TS alone were calculated per CS location and ISI 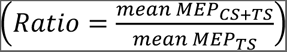. Ratios > 1 indicate facilitation of M1 output, while values < 1 indicate inhibition.

### 2.8. Statistical analysis

Statistical analysis was performed using JMP® (v16.2.0, SAS Institute Inc., Cary, NC, USA) and RStudio® (v2023.09.1+494, Posit Software, PBC, Boston, MA, USA). Normality assumptions for data were tested using the Shapiro-Wilk test, normality of residuals was assessed by visual inspection of the quantile-quantile (Q-Q) plots prior to further analyses. Assumptions of normality of the residuals were met for all conditions except ISI_PMd–M1_ of 10ms (see Supplement 1). For all statistical tests, the significance level was set to α = 0.05.

#### Neurophysiological measures

Paired-samples t-tests and Wilcoxon signed-ranks test were performed to examine potential differences in rMT, CS, and TS intensities between both measurement days for normally and non-normally distributed data, respectively.

#### Modulation of M1 output

To determine what factors significantly influence the modulation of M1 output, a full mixed model of MEP ratio was computed containing SESSION [day 1 vs. day 2], CS LOCATION [PMd vs. M1], and ISI [3,5,6,7,8,10 ms] as fixed effects and SUBJECT as random effects. This showed that SESSION did not significantly contribute to the model and was therefore removed. The final model was used to determine the effect of ISIs and CS LOCATION on the modulation of M1 with fixed effects of CS LOCATION and ISI, and the random effect SUBJECT. To determine if each condition had a significant difference from no modulation, we calculated the z-score using the standard error and estimated mean from our mixed model. Next, p-values were computed using the p-norm distribution function in RStudio. Post hoc pairwise comparison with Bonferroni correction was used to identify differences between conditions showing significant facilitation or inhibition. Additionally, the influence of head circumference and nasion-inion distance were tested for each ISI of the PMd–M1 condition using Bonferroni-corrected linear and quadratic regression analysis to check for a systematic influence of head size on M1 output modulation, that might be related to the fixed PMd–M1 stimulation distance.

#### Reliability

To evaluate the relative 24-hour test-retest reliability a mixed-effect model was implemented. The ICC was calculated as the ratio of subject variance and total variance of the mixed model, 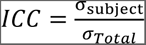 (Koo and Li, 2016). The SUBJECT term was entered into the model as a random effect to account for individual differences and variations within the study sample, while SESSION was entered as a fixed effect. To determine the significance of the ICC for a particular condition, we conducted a likelihood ratio test between the loglikelihood *L* of two nested mixed models *m1* and *m2*: 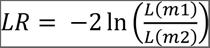. Here, *m1* refers to the full mixed model, while *m2* refers to a similar model but without the random SUBJECT effect. We computed the p-values in RStudio based on the chi-squared distribution. Lastly, the ICC values for the relative reliability were classified as follows: values between 0 to 0.39 indicated poor test-retest reliability, values between 0.40 to 0.59 indicated fair test-retest reliability, values between 0.60 to 0.74 indicated good test-retest reliability, and values between 0.75 to 1.00 indicated excellent test-retest reliability (Cicchetti, 1994).

Additionally, absolute reliability metrics such as the standard error of measurement (SEMeas), minimal detectable change (MDC), and coefficient of variation (CV) can indicate whether or not a chosen TMS outcome is a suitable and consistent biomarker for future research (Beaulieu et al., 2017, Beckerman et al., 2001, Weir, 2005). SEMeas was computed as 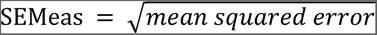 (Houde et al., 2018, Rurak et al., 2021, Schambra et al., 2015, Turco et al., 2019, Weir, 2005), and provides an indication of the range of scores that can be expected when retesting (Portney and Watkins, 2017). Hence, the smaller the SEMeas, the less variable and more reliable the measurement is, which in turn allows for more consistency in comparison across studies since it is not affected by between-subject variability, such as the ICC (Schambra et al., 2015, Weir, 2005). To allow for comparison between conditions, the SEMeas was divided by the pooled mean from day 1 and day 2, and expressed as a percentage: 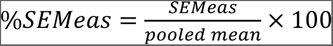 (Rurak et al., 2021, Turco et al., 2019). Additionally, the MDC was computed, which is the smallest amount of change an observation would need to show in order to exceed the random noise and become detectable. This was first done on the level of an individual by computing the 95% confidence interval (CI) of the SEMeas, i.e., 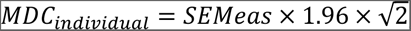, to then calculate the MDC on the group level using 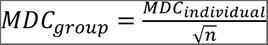 (Houde et al., 2018, Rurak et al., 2021, Schambra et al., 2015, Weir, 2005). If the change in the variable of interest exceeds the MDC, it is considered a true change with 95% confidence. Otherwise, it should be considered a measurement error. Therefore, a smaller MDC indicates a measure that is more sensitive to detect change. Lastly, the CV was computed as a measure of stability across repeated trials and represents the between-subject variability, with a smaller CV corresponding to a more reliable and consistent measure: 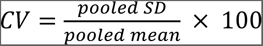 (Biabani et al., 2018, Houde et al., 2018, Rurak et al., 2021, Turco et al., 2019).

### 2.9. Methodological quality

The methodological quality of this study was assessed using the checklist proposed by Chipchase et al. (2012) for studies implementing TMS over M1 and scored 96.5% (see Supplement 2). The checklist contains 30 items probing the inclusion of correct and complete reporting information related to subjects (8 items), methodology (21 items), and analysis (2 items). Each item could be evaluated as ‘reported’ (i.e., described without further detail) or ‘controlled’ [e.g., excluded, used as a covariate in the analysis, or reported in sufficient (statistical) detail; see Beaulieu et al. (2017)]. The final score was computed as: 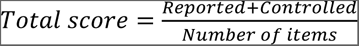 (Beaulieu et al., 2017).

## 3. RESULTS

### 3.1. Excluded datapoints

A small number of trials (5.76%) were excluded from the analysis for the following reasons: (1) stimulation was administered > 5 mm away from the TS or CS stimulus target location, as indicated by the neuronavigational software (0.16%); (2) EMG signals surpassed a root mean square of 20 μV in the period 100–50 ms before the TMS pulse in the left or right FDI (0.17%) (Cuypers et al., 2020, Hehl et al., 2020); (3) the TS-only condition was below 0.1 mV (ppTMS data of one participant: 0.65%); (4) the recorded EMG signal exceeded the maximum measurable amplitude of 10 mV of the EMG system (all data from one participant: 4.78%).

### 3.2. Comparison of TMS intensities day 1 vs. day 2

Comparison of TMS intensities of day 1 and 2 can be found in Table 2. Values are expressed as a percentage maximum stimulator output (%MSO). Paired samples t-tests showed no significant difference in rMT (mean difference = -0.81 %MSO, t(20) = -1.23, p = 0.232) and CS intensity (mean difference = -1.48 %MSO, t(20) = -1.45, p = 0.163) of the MC-B65HO coil between measurement days. Wilcoxon signed-ranks tests showed no significant difference between measurement days for the rMT (median difference = 0 %MSO, W = 47.00, p = 0.275) and TS intensity (median difference = 0 %MSO, W = 56.50, p = 0.354) of the MC-B70 coil (see Supplement 3 for visualization using boxplots).

**Table 2.**
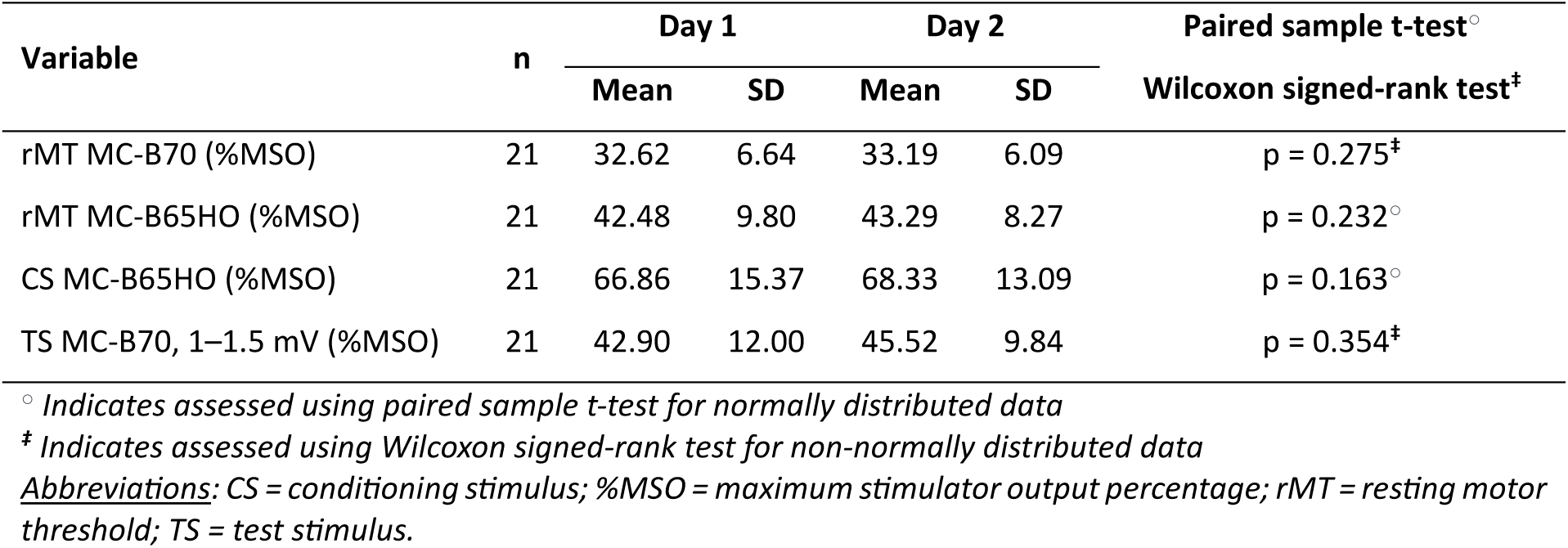
Comparison of TMS intensities day 1 vs. day 2.

### 3.3. Modulation of M1

A significant effect of ISI (F_5, 291.5_ = 71.901, p < 0.001) and CS target location (F_1, 292.1_ = 5.62, p = 0.018) on M1 output was found. Significant inhibition of M1 output occurred at an ISI_PMd–M1_ 3 ms (β = 0.47, SD = 0.18, 95% CI [0.39–0.55], p < 0.001), ISI_PMd–M1_ 5 ms (β = 0.80, SD = 0.20, 95% CI [0.72–0.89], p < 0.001) and SICI_3ms_ (β = 0.38, SD = 0.17, 95% CI [0.30, 0.46], p < 0.001). Significant facilitation of M1 output occurred at an ISI_PMd–M1_ 10 ms (β = 1.19, SD = 0.18, 95% CI [1.11–1.27], p < 0.001) and ICF_10ms_ (β = 1.10, SD = 0.17, 95% CI [0.02–1.17], p = 0.017) (see Table 3 and Figure 2).

**Figure 2.**
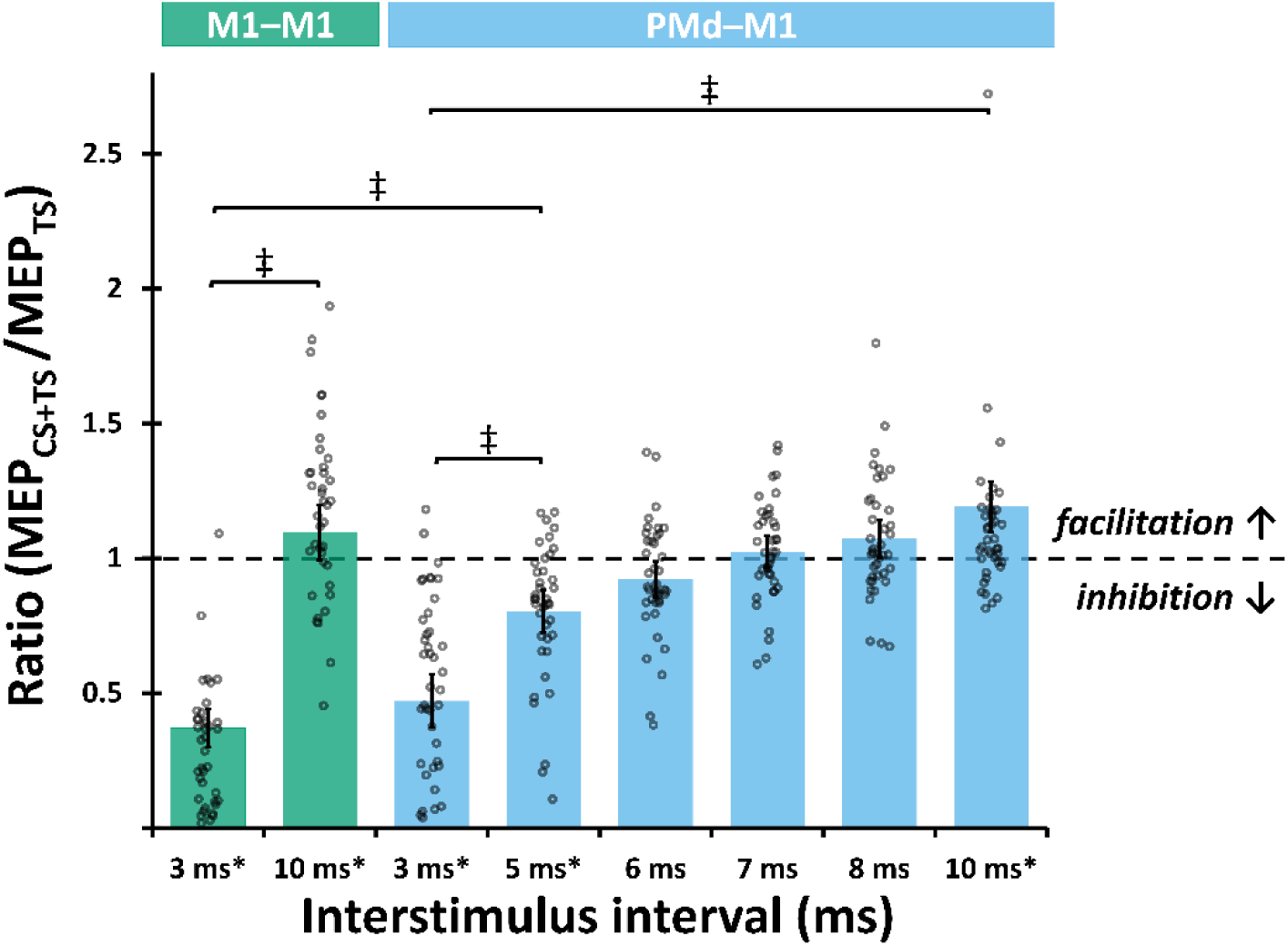
Effect of ISIs and CS target locations on M1 output and post hoc comparison between significantly modulated conditions. Asterisks indicate significant modulation of M1 output (p < 0.05). ‡ indicates significant differences between conditions (p < 0.01, Bonferroni corrected). Abbreviations: CS = conditioning stimulus; TS = test stimulus; M1 = primary motor cortex; PMd = dorsal premotor cortex.

**Table 3.**
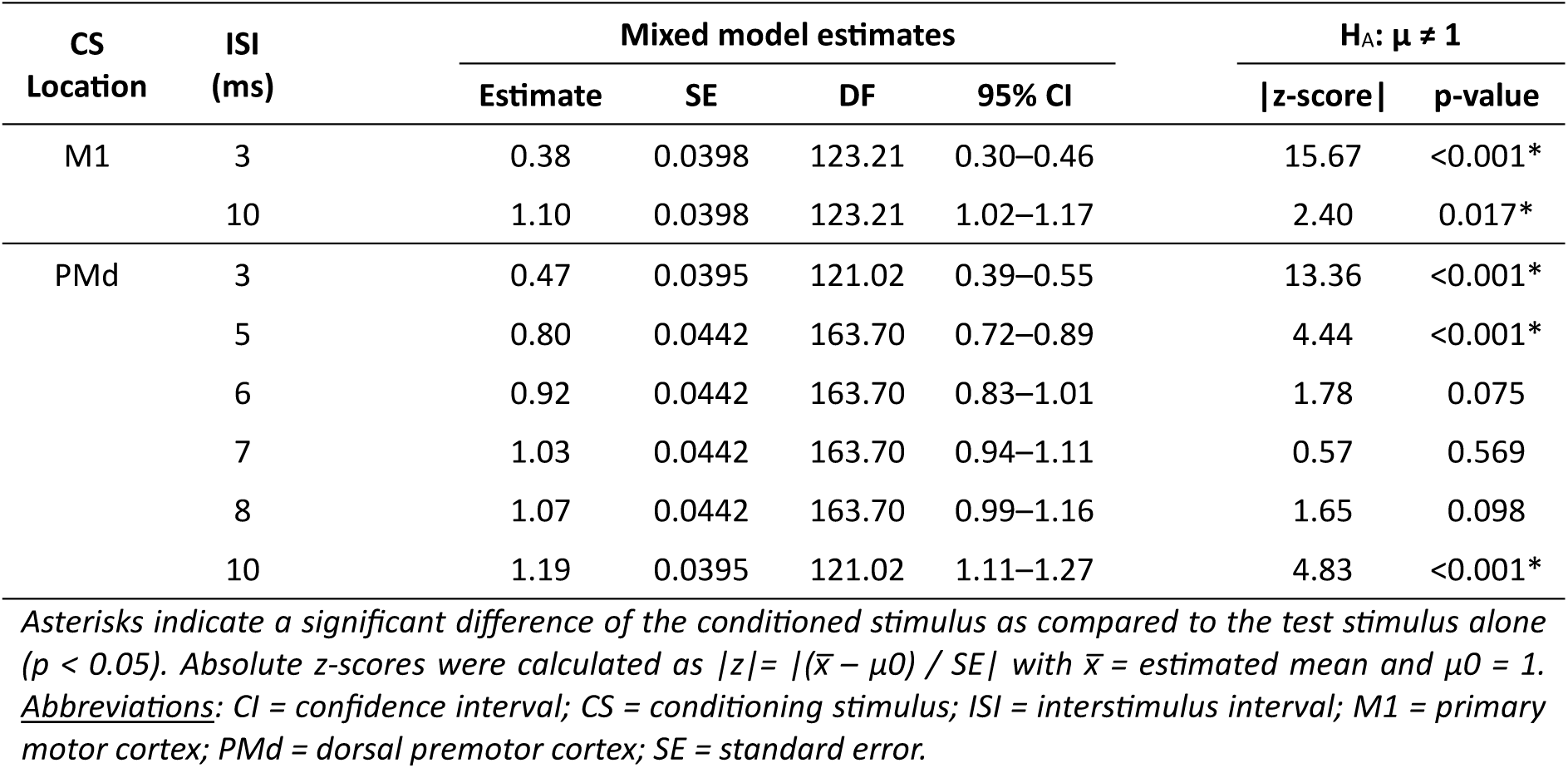
Effect of ISIs and CS target locations on M1 output.

Post hoc multiple pairwise comparisons of conditions that significantly modulated M1 output (with Bonferroni correction for 5 comparisons, i.e., α = 0.05/5) revealed that ISI_PMd–M1_ 3 ms vs. ISI_PMd–M1_ 10 ms, as well as SICI_3m_ vs. ICF_10ms_ resulted in significantly different modulation effects (i.e., inhibition and facilitation respectively for both; ISI_PMd–M1_ 3 ms vs. ISI_PMd–M1_ 10 ms: t(291.7) = -17.88, p < 0.001; SICI_3m_ vs. ICF_10ms_: t(291.7) = -17.88, p < 0.001). Furthermore, both ISI_PMd–M1_ 3 ms and SICI_3ms_ were both found to be significantly more inhibitory compared to ISI_PMd–M1_ 5 ms (ISI_PMd–M1_ 3 ms vs. 5ms: t(291.7) = -6.27, p < 0.001; SICI_3ms_ vs. ISI_PMd–M1_ 5 ms: t(291.7) = -8.04, p < 0.001), but were not statistically different from each other (ISI_PMd–M1_ 3 ms vs. SICI_3ms_: t(291.7) = -2.37, p = 0.0184) (see Figure 2). Head size (circumference and nasion-inion distance) had no significant influence on M1 output (all, p > 0.35).

### 3.4. Test-retest reliability

#### Relative reliability

Excellent test-retest reliability was found for SICI_3m_ (ICC = 0.81, p < 0.001), fair test-retest reliability was found for ISI_PMd–M1_ 3 ms (ICC = 0.47, p = 0.026), and ISI_PMd–M1_ 6 ms (ICC = 0.44, p = 0.038). All other conditions showed no ICC values that were significantly different from 0 and yielded poor test-retest reliability results (see Table 4).

**Table 4.** Relative and absolute 24-hour test-retest reliability.

| CS Location | ISI (ms) | n | Relative Reliability |  | Absolute reliability |  |  | CV (%) |
| --- | --- | --- | --- | --- | --- | --- | --- | --- |
|  |  |  | ICC | p-value | SEMeas (%) | MDC<sub>ind</sub> | MDC<sub>group</sub> |  |
| M1 | 3 | 19 | 0.8108 | 0.000* | 0.14 (46.02) | 0.39 | 0.09 | 75.13 |
|  | 10 | 19 | 0.2486 | 0.293 | 0.39 (32.63) | 1.08 | 0.25 | 26.66 |
| PMd | 3 | 20 | 0.4675 | 0.026* | 0.33 (59.98) | 0.91 | 0.20 | 58.41 |
|  | 5 | 20 | 0.3105 | 0.154 | 0.30 (37.13) | 0.83 | 0.18 | 31.48 |
|  | 6 | 20 | 0.4410 | 0.038* | 0.22 (24.26) | 0.62 | 0.14 | 23.23 |
|  | 7 | 20 | -0.2138 | 0.333 | 0.29 (28.28) | 0.80 | 0.18 | 18.38 |
|  | 8 | 20 | 0.2411 | 0.274 | 0.27 (25.44) | 0.76 | 0.17 | 20.88 |
|  | 10 | 20 | 0.1118 | 0.616 | 0.40 (36.07) | 1.12 | 0.25 | 27.11 |

Asterisks indicate intra-class correlation coefficients (ICCs) that are significantly different from 0 ( $p < 0.05$ ).
**Abbreviations:** CV = coefficient of variation; ICC = intraclass correlation coefficient; ISI = interstimulus interval; MDC<sub>ind</sub> = minimal detectable change for individuals; MDC<sub>group</sub> = minimal detectable change for groups; SEMeas = standard error of the measurement.

#### Absolute reliability

The highest %SEMeas values were observed for ISI_PMd–M1_ 3 ms and SICI_3ms_, while the lowest %SEMeas was observed at an ISI_PMd–M1_ of 6 ms, 7 ms, and 8 ms (see Table 4). MDC scores ranged from 0.39 (ISI_PMd–M1_ of 10 ms) to 1.12 (ICF_10ms_), suggesting that changes exceeding these values can be considered significant beyond random variation. However, MDC_group_ (using a sample size of 19 for CS location M1 and 20 for CS location PMd) was below 0.25 for all conditions. Stability across repeated trails was highest for ISI_PMd–M1_ at 3 ms and SICI_3ms_ whereas ISI_PMd–M1_ at 6, 7, and 8 ms showed the lowest spread, as reflected by their respective CV (see Table 4).

## 4. DISCUSSION

The present study revealed three main findings. First, we explored the modulatory effect of various ISIs using a stacked dsTMS setup for probing intrahemispheric PMd–M1 interactions and found a significant inhibitory effect at ISIs of 3 and 5 ms, as well as a significant facilitatory effect when using a 10 ms ISI. Second, investigating the 24-hour test-retest reliability of the current dsTMS set-up and protocol yielded fair test-retest reliability when testing the intrahemispheric PMd–M1 interaction with ISIs of 3 and 6 ms. Third, we validated the concept of this dsTMS setup by comparing ppTMS outcomes such as SICI and ICF to well-established values in the literature and showed reproducibility of classic ppTMS protocols as well as excellent test-retest reliability for SICI measurements.

### 4.1. Modulation of M1 output when conditioning ipsilateral PMd and comparison to paired-pulse protocols

Well-coordinated co-activation of M1 with other parts of the supplementary motor system is required to produce successful movement (Rodriguez-Nieto et al., 2022). More specifically, M1 is assumed to serve as a convergence point for input of several nodes of the motor system, including PMd. This functional dependency between M1 and PMd for essentially any movement suggests the presence of a close functional, structural, and effective connectivity (Babaeeghazvini et al., 2019, Bencivenga et al., 2021, Dirren et al., 2021, Lefebvre et al., 2017, McGregor and Gribble, 2017). dsTMS offers an option to measure this effective connectivity at rest and during task performance, providing information about the physiology and timing of the PMd–M1 connection.

When stimulating M1 with TMS, distinct descending volleys can be measured in the corticospinal neurons and will summate at the level of the spinal cord to finally elicit an MEP in the muscle corresponding to the stimulated M1 area. When stimulating at higher intensities, this results in direct activation of the corticospinal neurons (i.e., D-wave), whereas lower intensities rather elicit an indirect activation of M1 via mono- and polysynaptic inputs of the local intracortical network (I-waves), to which PMd contributes (Di Lazzaro et al., 2001, Groppa et al., 2012, Liao et al., 2023, Rossini et al., 2015, Ziemann et al., 1996). Based on the emerging I-waves, choices in the dsTMS stimulation protocol such as the intensity of the CS and TS applied to PMd and M1 respectively, their timing (ISI), and the coil orientation (Di Lazzaro et al., 2001) can influence the pattern of activation in M1 and the subsequent muscle response.

In the present study, conditioning PMd resulted in a strong inhibition of M1 output at an ISI of 3 and 5 ms. This is in line with a recent study which implemented the same stacked dsTMS setup (Hehl et al., 2024) that showed strong inhibition of M1 output at an ISI of 3 ms. To date, to the best of our knowledge no other studies investigated the effect of intrahemispheric PMd–M1 stimulation at an ISI of 5 ms.

In contrast, the present study did not yield a significant modulation of M1 output at an ISI 6 ms. Previous research yielded inconsistent results, including moderate inhibition of M1 output (Civardi et al., 2001, Parmigiani et al., 2015, Parmigiani et al., 2018), a modulatory effect that was highly individual and could either be inhibitory or facilitatory (Hehl et al., 2024), as well as no modulatory effect (Bäumer et al., 2009, Van Hoornweder et al., 2021). Variable study designs (stimulation intensities, placement of CS coil, coil focality, etc.) might explain these diverse results (Van Malderen et al., 2023). Furthermore, this study encompassed 20 datapoints in the final analysis. When entering the obtained results for the PMd–M1 interaction with an ISI of 6 ms into a power calculation (Cohen’s *d* = 0.4 based on reported results, α = 0.05), this study would have needed to include 41 participants to obtain a power (1-β) of 0.8. Indeed, when performing an analysis on the combined dataset of Hehl et al. (2024) and the day 1 data of the present study that were collected with the same methodology, the PMd–M1 interaction with an ISI of 6 ms showed a significant inhibition (n = 43, mean MEP ratio = 0.912 ± 0.278 SD, 95% CI [0.826–0.998], t(42) = -2.076, p = 0.022 [one-sided, H_0_: 1 ≥ t]). This means, that bigger sample sizes and/or studies investigating intra-individual differences between conditions (i.e., rest vs. movement preparation) might be needed to increase study power.

PMd–M1 stimulation at an ISI of 7 ms in the present study yielded no significant modulatory effect on M1 output, and so far, no reports investigated this ISI in a similar context. Likewise, stimulation at an ISI of 8 ms did not modify M1 output and this absence of modulation was confirmed by other studies (Civardi et al., 2001, Parmigiani et al., 2015, Parmigiani et al., 2018, Vesia et al., 2017). Only one sub-study by Bäumer et al. (2009) including eight participants yielded a significant inhibitory effect of an 8 ms ISI when stimulating the PMd as close to M1 as technically possible with an intensity of 90% aMT.

Lastly, facilitation of M1 output was elicited when stimulating at an ISI of 10 ms. This is in contradiction with previous research that reports no significant modulatory at an ISI of 10 ms (Bäumer et al., 2009, Civardi et al., 2001). Again, diversity in study design and coil placement might explain these discrepancies.

To further support the concept of the stacked dsTMS setup, we additionally investigated the ability to replicate well-investigated conventional single-coil paired-pulse paradigms such as SICI and ICF. In line with extensive previous research (Biabani et al., 2018, Boroojerdi et al., 2000, Cuypers et al., 2021, Du and Hong, 2018, Hermans et al., 2019, Ngomo et al., 2012, Nielsen et al., 2021, Samusyte et al., 2018, Vaalto et al., 2011, Wessel et al., 2019) the current sample revealed strong (62%) inhibition when conditioning M1 at an ISI of 3 ms. Likewise, the modulation of M1 output observed following an

ICF protocol with an ISI of 10 ms resulted in 10% facilitation, falling in line with previous findings (Kujirai et al., 1993, Wassermann, 2002). This confirms that the stacked dsTMS set-up can elicit comparable effects as observed with classical single coil paradigms, which will be further supported by the excellent reliability of the SICI measurement discussed below.

Finally, we compared conditions significantly modulating the M1 output with each other. A comparison between the SICI condition and the PMd–M1 interaction tested with the same 3 ms ISI yielded no significant difference in the amount of inhibition. When comparing the inhibitory PMd–M1 interactions, we found that stimulation with an ISI of 3 ms was more inhibitory than stimulation at an ISI of 5 ms (mean inhibition of 53% vs. 20% respectively). Although the ISIs between 6 and 8 ms where not significantly modulating M1 output, less inhibition with increasing ISIs can be observed, with an ISI of 10 ms significantly facilitating M1 and showing no significant difference from the facilitation observed with the ICF paradigm. In summary, notably, the PMd–M1 interaction measures in this study, but also in the literature, followed a similar temporal relationship as compared to SICI measurements. More specifically, inhibition has been reported for ISIs up to 4 or 5 ms (Fong et al., 2021, Nielsen et al., 2021, Tankisi et al., 2021, Ziemann et al., 1996) but not for 6 and 7 ms (Du et al., 2014, Nielsen et al., 2021, Tankisi et al., 2021, Ziemann et al., 1996). This might point towards one of the two following possible explanations. On the one hand, the conditioning pulse of PMd might have co-activated M1 in a way that elicits very similar processes as compared to SICI and ICF measurements, i.e., intrahemispheric M1–M1 interactions. However, a single-case multi-locus TMS study points towards an insufficient current overflow for CS–TS distances of more than 2 cm (and 0.8 cm specifically for SICI) (Nieminen et al., 2022). Consequently, the PMd–M1 stimulation distance of 2.1 cm implemented in the present study is a counter-argument for a mechanism similar to SICI and ICF measurements due to current overflow. However, to date, comparability studies between multi-locus TMS and standard TMS coils are scarce and the bigger coil size in the present study might influence the results due to more current spread. On the other hand, PMd–M1 interactions might simply be organized in a very similar way to M1–M1 interactions based on their close anatomic (Picard and Strick, 2001) and functional (Volz et al., 2015) relationship. Unfortunately, the present study design does not provide sufficient results to challenge either of these hypotheses.

### 4.2. 24-hour test-retest reliability

The second aim of this study was to establish the 24-hour test-retest reliability following PMd–M1 stimulation at the various implemented ISIs. For the PMd–M1 interaction measures, fair relative test-retest reliability was observed at ISIs of 3 ms (ICC: 0.47) and 6 ms (ICC: 0.44). However, even though scoring similarly on relative reliability, data spread (i.e., MDC and SEMeas), and variability around the measured mean (i.e., CV) were found to be larger in the ISI 3 ms as compared to the 6 ms condition. Furthermore, in the conditions inhibiting M1 output, the inter-individual variability (i.e., CV) seems to decrease as the influence of the inhibitory circuitry in M1 decreases, along with the observed error estimate (i.e., SEMeas).

Results obtained of PMd–M1 stimulation at ISI of 6 and 8 ms (ICC: 0.44 and 0.24 respectively) were comparable with previous dsTMS literature probing SMA–M1 connectivity, which reported an ICC of 0.40 (ISI 6 ms) and 0.20 (ISI 8 ms) (Rurak et al., 2021). However, the present study yielded no test-retest reliability for an ISI of 7 ms, whereas an ICC of 0.69 was reported for the SMA–M1 interaction (Rurak et al., 2021). When comparing the absolute reliability metrics to those of previous studies, PMd–M1 and SMA–M1 stimulation at an ISI of 6 ms showed similar within- and between-subject variance (i.e., SEMeas and CV, respectively). In contrast, ISIs of 7 and 8 ms showed greater within- and between-subject variance in the present PMd–M1 study and, as a result, larger individual and group MDC indexes than for the SMA–M1 connectivity (Rurak et al., 2021).

Additionally, the obtained reliability results of SICI and ICF of the present study were found to be comparable to values of studies using conventional one-coil paired-pulse SICI and ICF paradigms. SICI showed excellent test-retest reliability with an ICC of 0.81, in line with previous literature reporting ICCs ranging from 0.23 to 0.91 (Biabani et al., 2018, Du et al., 2014, Fleming et al., 2012, Hermsen et al., 2016, Matamala et al., 2018, Ngomo et al., 2012). While not commonly reported, the inter-individual variability (CV) in our sample was 75%, and thus remarkably higher than the inter-individual variability in other conditions. However, the calculation of this measure needs to be considered (ratio between pooled SD and pooled mean) since it can be inflated specifically for measures with a low pooled mean such as the strongly inhibiting 3 ms M1–M1 and PMd–M1 conditions, despite or rather because of a similar SD with other ISI conditions (Biabani et al., 2018). For ICF, poor non-significant relative reliability with an ICC of 0.11 was observed in the present study, which is in line with previous research with ICCs ranging from 0 to 0.45 (Biabani et al., 2018, Hermsen et al., 2016). In contrast to SICI, the inter-individual variability (CV) for ICF was lower (27%), which might simply be explained by the higher mean MEP ratio for ICF as compared to SICI but a similar SD. Lastly, caution is needed when interpreting and comparing reliability metrics across studies since most did not report between-subject variance or confidence intervals, which can inflate ICCs (Schambra et al., 2015, Weir, 2005).

Individual MDCs were high in all stimulation conditions ranging from 67–166% of the respectively measured mean, indicating significant changes in MEP amplitude or connectivity would be required to consider the change in a single participant genuine with 95% confidence. Therefore, the use of these metrics as biomarkers on an individual level is not advised. However, when accounting for the sample size, MDC metrics decrease (range 15–37% of the respectively pooled mean) making them more suitable for the detection of group-level changes such as implemented in most studies. Since MDC values are not commonly reported in practice, no values could be found for comparison of the PMd–M1 results. However, MDCs for SICI and ICF measures have been more commonly reported, ranging from 17–60% for SICI and 59–100% for ICF (Beaulieu et al., 2017, Ngomo et al., 2012, Schambra et al., 2015). For SICI (29%) our results are in line with the reported MDC_group_ values. In contrast, the MDC_group_ for ICF (21%) in the current study is much lower than the range reported previously.

### 4.3. Limitations and future perspectives

Even though the current study scored excellent (96%) on the methodological quality checklist of Chipchase et al. (2012) a few limitations and considerations for future research should be considered. First, the investigated study sample consisted of healthy, right-handed, young adults. Therefore generalizability to other populations might be limited since reliability metrics can most confidently be compared between similar study samples (Houde et al., 2018, Rurak et al., 2021, Schambra et al., 2015, Weir, 2005), and effective PMd–M1 connectivity of, e.g., older adults might differ significantly from the present sample due to connectivity declines with increasing age (Ni et al., 2009), and/or show a higher variability.

Secondly, the present study only investigated modulatory PMd–M1 effects at rest, but not during task preparation or execution. Previous research has shown both facilitatory and inhibitory PMd–M1 interactions at rest. However, responses to stimulation during motor tasks seem to lean more towards facilitation, whereas inhibition seemed to be more pronounced in the context of action withholding (Van Malderen et al., 2023). Hence, it is not possible to predict whether the investigated ISI in task-based protocols would yield comparable results in terms of modulation and reliability.

Third, only 24-hour test-retest reliability was investigated in the present study. Therefore, we cannot draw any conclusions about the test-retest reliability within the same session or across larger time spans, which would necessitate another experimental design (Matamala et al., 2018, O’Leary et al., 2015, Pellegrini et al., 2018, Peri et al., 2017).

Lastly, to ensure that the experiment could be completed without major fatigue effects on the participant, a limited number of conditions such as ISIs, intensities, current directions, and CS locations could be evaluated. Future research might elaborate other conditions and provide a more complete picture of the effective intrahemispheric PMd–M1 interaction at rest and during motor tasks.

## 5. CONCLUSION

Based on the present study, we can confirm the ability of the stacked dsTMS set-up to probe both inhibitory (ISI = 3 and 5 ms) and facilitatory (ISI = 10 ms) PMd–M1 interactions, with a fair 24-hour test-retest reliability for ISIs of 3 and 6 ms. Furthermore, we confirmed the ability of this setup to replicate conventional SICI and ICF paradigm outcomes with excellent test-retest reliability for SICI, which further supports the concept of the stacked dsTMS setup to probe intrahemispheric interactions. Furthermore, the minimal detectable change results point towards usability of the intrahemispheric PMd–M1 measures for studies investigating changes on the group level (i.e., in a research context), but not for an individual (i.e., in a clinical context). Due to the importance of the PMd during motor function, future research should focus on further disentangling the intricate relationship between M1 and ipsilateral PMd by further optimizing and investigating study protocols and the underlying reliability metrics. This is of importance since investigating intrahemispheric PMd–M1 effective connectivity offers promising possibilities to better understand motor control and movement disorders.

## Data Availability

The data that support the findings of this study are available from the corresponding author upon reasonable request.

## AUTHOR CONTRIBUTIONS

**Robin Heemels:** Conceptualization; Formal Analysis; Investigation; Methodology; Project Administration; Visualization; Writing – Original Draft Preparation; Writing – Review & Editing. **Sian Ademi:** Conceptualization; Formal Analysis; Investigation; Methodology; Project Administration; Writing – Original Draft Preparation. **Melina Hehl:** Conceptualization; Investigation; Methodology; Data Curation; Funding Acquisition; Resources; Software; Supervision; Writing – Review & Editing.

## ACKNOWLEDGEMENTS

This work was supported by the Research Fund KU Leuven (C16/15/070), the Research Foundation Flanders grant (G039821N), the Excellence of Science grant (EOS 30446199, MEMODYN) and the Hercules fund AUHL/11/01 (R-3987) and I005018N. Melina Hehl (11F6921N) is funded by a fellowship grant from Research Foundation Flanders. The authors declare no competing financial interests. The funders had no role in study design, data collection and analysis, decision to publish, or preparation of the manuscript.

## ABBREVIATIONS

aMT: active motor threshold
CI: confidence interval
CS: conditioning stimulus
CV: coefficient of variation
dsTMS: dual-site transcranial magnetic stimulation
EMG: electromyography
FDI: first dorsal interosseus
ICC: intra-class correlation coefficients
ICF: intracortical facilitation
ISI: interstimulus interval
M1: primary motor cortex
MDC: minimal detectable change
MEP: motor-evoked potentials
MSO: maximum stimulator output
PMd: dorsal premotor cortex
ppTMS: paired-pulse TMS
rMT: resting motor threshold
SEMeas: standard error of measurement
SICI: short-interval intracortical inhibition
SMA: supplementary motor area
TMS: transcranial magnetic stimulation
TS: test stimulus

## REFERENCES

Abe M, Hanakawa T. Functional coupling underlying motor and cognitive functions of the dorsal premotor cortex. Behav Brain Res 2009;198(1):13–23. doi: 10.1016/j.bbr.2008.10.046

Amiez C, Kostopoulos P, Champod AS, Petrides M. Local morphology predicts functional organization of the dorsal premotor region in the human brain. J Neurosci 2006;26(10):2724–31. doi: 10.1523/JNEUROSCI.4739-05.2006

Babaeeghazvini P, Rueda-Delgado LM, Zivari Adab H, Gooijers J, Swinnen S, Daffertshofer A. A combined diffusion-weighted and electroencephalography study on age-related differences in connectivity in the motor network during bimanual performance. Hum Brain Mapp 2019;40(6):1799–813. doi: 10.1002/hbm.24491

Bäumer T, Schippling S, Kroeger J, Zittel S, Koch G, Thomalla G, et al. Inhibitory and facilitatory connectivity from ventral premotor to primary motor cortex in healthy humans at rest--a bifocal TMS study. Clin Neurophysiol 2009;120(9):1724–31. doi: 10.1016/j.clinph.2009.07.035

Beaulieu LD, Flamand VH, Masse-Alarie H, Schneider C. Reliability and minimal detectable change of transcranial magnetic stimulation outcomes in healthy adults: A systematic review. Brain Stimul 2017;10(2):196–213. doi: 10.1016/j.brs.2016.12.008

Beck AT, Steer RA, Ball R, Ranieri W. Comparison of Beck Depression Inventories -IA and -II in psychiatric outpatients. J Pers Assess 1996;67(3):588–97. doi: 10.1207/s15327752jpa6703_13

Beckerman H, Roebroeck ME, Lankhorst GJ, Becher JG, Bezemer PD, Verbeek AL. Smallest real difference, a link between reproducibility and responsiveness. Qual Life Res 2001;10(7):571–8. doi: 10.1023/a:1013138911638

Beets IA, Gooijers J, Boisgontier MP, Pauwels L, Coxon JP, Wittenberg G, et al. Reduced Neural Differentiation Between Feedback Conditions After Bimanual Coordination Training with and without Augmented Visual Feedback. Cereb Cortex 2015;25(7):1958–69. doi: 10.1093/cercor/bhu005

Bencivenga F, Sulpizio V, Tullo MG, Galati G. Assessing the effective connectivity of premotor areas during real vs imagined grasping: a DCM-PEB approach. NeuroImage 2021;230:117806. doi: 10.1016/j.neuroimage.2021.117806

Biabani M, Farrell M, Zoghi M, Egan G, Jaberzadeh S. The minimal number of TMS trials required for the reliable assessment of corticospinal excitability, short interval intracortical inhibition, and intracortical facilitation. Neurosci Lett 2018;674:94–100. doi: 10.1016/j.neulet.2018.03.026

Boroojerdi B, Kopylev L, Battaglia F, Facchini S, Ziemann U, Muellbacher W, et al. Reproducibility of intracortical inhibition and facilitation using the paired-pulse paradigm. Muscle Nerve 2000;23(10):1594–7. doi: 10.1002/1097-4598(200010)23:10<1594::aid-mus19>3.0.co;2-3

Chipchase L, Schabrun S, Cohen L, Hodges P, Ridding M, Rothwell J, et al. A checklist for assessing the methodological quality of studies using transcranial magnetic stimulation to study the motor system: an international consensus study. Clin Neurophysiol 2012;123(9):1698–704. doi: 10.1016/j.clinph.2012.05.003

Cicchetti D. Guidelines, Criteria, and Rules of Thumb for Evaluating Normed and Standardized Assessment Instrument in Psychology. Psychological Assessment 1994;6:284–90. doi: 10.1037/1040-3590.6.4.284

Civardi C, Cantello R, Asselman P, Rothwell JC. Transcranial magnetic stimulation can be used to test connections to primary motor areas from frontal and medial cortex in humans. NeuroImage 2001;14(6):1444–53. doi: 10.1006/nimg.2001.0918

Cuypers K, Hehl M, van Aalst J, Chalavi S, Mikkelsen M, Van Laere K, et al. Age-related GABAergic differences in the primary sensorimotor cortex: A multimodal approach combining PET, MRS and TMS. NeuroImage 2021;226:117536. doi: 10.1016/j.neuroimage.2020.117536

Cuypers K, Verstraelen S, Maes C, Hermans L, Hehl M, Heise KF, et al. Task-related measures of short-interval intracortical inhibition and GABA levels in healthy young and older adults: A multimodal TMS-MRS study. NeuroImage 2020;208:116470. doi: 10.1016/j.neuroimage.2019.116470

Di Lazzaro V, Oliviero A, Mazzone P, Insola A, Pilato F, Saturno E, et al. Comparison of descending volleys evoked by monophasic and biphasic magnetic stimulation of the motor cortex in conscious humans. Exp Brain Res 2001;141(1):121–7. doi: 10.1007/s002210100863

Dirren E, Bourgeois A, Klug J, Kleinschmidt A, van Assche M, Carrera E. The neural correlates of intermanual transfer. NeuroImage 2021;245:118657. doi: 10.1016/j.neuroimage.2021.118657

Du X, Hong LE. Test-retest reliability of short-interval intracortical inhibition and intracortical facilitation in patients with schizophrenia. Psychiatry Res 2018;267:575–81. doi: 10.1016/j.psychres.2018.06.014

Du X, Summerfelt A, Chiappelli J, Holcomb HH, Hong LE. Individualized brain inhibition and excitation profile in response to paired-pulse TMS. J Mot Behav 2014;46(1):39–48. doi: 10.1080/00222895.2013.850401

Fiori F, Chiappini E, Soriano M, Paracampo R, Romei V, Borgomaneri S, et al. Long-latency modulation of motor cortex excitability by ipsilateral posterior inferior frontal gyrus and pre-supplementary motor area. Sci Rep 2016;6:38396. doi: 10.1038/srep38396

Fleming MK, Sorinola IO, Newham DJ, Roberts-Lewis SF, Bergmann JH. The effect of coil type and navigation on the reliability of transcranial magnetic stimulation. IEEE Trans Neural Syst Rehabil Eng 2012;20(5):617–25. doi: 10.1109/TNSRE.2012.2202692

Fong PY, Spampinato D, Rocchi L, Hannah R, Teng Y, Di Santo A, et al. Two forms of short-interval intracortical inhibition in human motor cortex. Brain Stimul 2021;14(5):1340–52. doi: 10.1016/j.brs.2021.08.022

Fukushima M, Betzel RF, He Y, van den Heuvel MP, Zuo XN, Sporns O. Structure-function relationships during segregated and integrated network states of human brain functional connectivity. Brain Struct Funct 2018;223(3):1091–106. doi: 10.1007/s00429-017-1539-3

Grafton ST, Fagg AH, Arbib MA. Dorsal premotor cortex and conditional movement selection: A PET functional mapping study. J Neurophysiol 1998;79(2):1092–7. doi: 10.1152/jn.1998.79.2.1092

Groppa S, Werner-Petroll N, Munchau A, Deuschl G, Ruschworth MF, Siebner HR. A novel dual-site transcranial magnetic stimulation paradigm to probe fast facilitatory inputs from ipsilateral dorsal premotor cortex to primary motor cortex. NeuroImage 2012;62(1):500–9. doi: 10.1016/j.neuroimage.2012.05.023

Halsband U, Passingham RE. Premotor cortex and the conditions for movement in monkeys (Macaca fascicularis). Behav Brain Res 1985;18(3):269–77. doi: 10.1016/0166-4328(85)90035-x

Hehl M, Swinnen SP, Cuypers K. Alterations of hand sensorimotor function and cortical motor representations over the adult lifespan. Aging (Albany NY) 2020;12(5):4617–40. doi: 10.18632/aging.102925

Hehl M, Van Malderen S, Geraerts M, Meesen RLJ, Rothwell JC, Swinnen SP, et al. Probing intrahemispheric interactions with a novel dual-site TMS setup. Clin Neurophysiol 2024;158:180–95. doi: 10.1016/j.clinph.2023.12.128

Hermans L, Maes C, Pauwels L, Cuypers K, Heise KF, Swinnen SP, et al. Age-related alterations in the modulation of intracortical inhibition during stopping of actions. Aging (Albany NY) 2019;11(2):371–85. doi: 10.18632/aging.101741

Hermsen AM, Haag A, Duddek C, Balkenhol K, Bugiel H, Bauer S, et al. Test-retest reliability of single and paired pulse transcranial magnetic stimulation parameters in healthy subjects. J Neurol Sci 2016;362:209–16. doi: 10.1016/j.jns.2016.01.039

Houde F, Laroche S, Thivierge V, Martel M, Harvey MP, Daigle F, et al. Transcranial Magnetic Stimulation Measures in the Elderly: Reliability, Smallest Detectable Change and the Potential Influence of Lifestyle Habits. Front Aging Neurosci 2018;10:379. doi: 10.3389/fnagi.2018.00379

Klem GH, Luders HO, Jasper HH, Elger C. The ten-twenty electrode system of the International Federation. The International Federation of Clinical Neurophysiology. Electroencephalogr Clin Neurophysiol Suppl 1999;52:3–6. doi: 10.1080/00029238.1961.11080571

Koch G, Franca M, Del Olmo MF, Cheeran B, Milton R, Alvarez Sauco M, et al. Time course of functional connectivity between dorsal premotor and contralateral motor cortex during movement selection. J Neurosci 2006;26(28):7452–9. doi: 10.1523/JNEUROSCI.1158-06.2006

Koo TK, Li MY. A Guideline of Selecting and Reporting Intraclass Correlation Coefficients for Reliability Research. J Chiropr Med 2016;15(2):155–63. doi: 10.1016/j.jcm.2016.02.012

Kujirai T, Caramia MD, Rothwell JC, Day BL, Thompson PD, Ferbert A, et al. Corticocortical inhibition in human motor cortex. J Physiol 1993;471:501–19. doi: 10.1113/jphysiol.1993.sp019912

Lefebvre S, Dricot L, Laloux P, Desfontaines P, Evrard F, Peeters A, et al. Increased functional connectivity one week after motor learning and tDCS in stroke patients. Neuroscience 2017;340:424–35. doi: 10.1016/j.neuroscience.2016.10.066

Li N, Chen TW, Guo ZV, Gerfen CR, Svoboda K. A motor cortex circuit for motor planning and movement. Nature 2015;519(7541):51-6. doi: 10.1038/nature14178

Liao WY, Opie GM, Ziemann U, Semmler JG. Modulation of dorsal premotor cortex differentially influences I-wave excitability in primary motor cortex of young and older adults. J Physiol 2023;601(14):2959–74. doi: 10.1113/JP284204

Ly JQM, Gaggioni G, Chellappa SL, Papachilleos S, Brzozowski A, Borsu C, et al. Circadian regulation of human cortical excitability. Nat Commun 2016;7:11828. doi: 10.1038/ncomms11828

Matamala JM, Howells J, Dharmadasa T, Trinh T, Ma Y, Lera L, et al. Inter-session reliability of short-interval intracortical inhibition measured by threshold tracking TMS. Neurosci Lett 2018;674:18–23. doi: 10.1016/j.neulet.2018.02.065

McGregor HR, Gribble PL. Functional connectivity between somatosensory and motor brain areas predicts individual differences in motor learning by observing. J Neurophysiol 2017;118(2):1235–43. doi: 10.1152/jn.00275.2017

Nakayama Y, Yamagata T, Tanji J, Hoshi E. Transformation of a virtual action plan into a motor plan in the premotor cortex. J Neurosci 2008;28(41):10287–97. doi: 10.1523/JNEUROSCI.2372-08.2008

Nasreddine ZS, Phillips NA, Bedirian V, Charbonneau S, Whitehead V, Collin I, et al. The Montreal Cognitive Assessment, MoCA: a brief screening tool for mild cognitive impairment. J Am Geriatr Soc 2005;53(4):695–9. doi: 10.1111/j.1532-5415.2005.53221.x

Ngomo S, Leonard G, Moffet H, Mercier C. Comparison of transcranial magnetic stimulation measures obtained at rest and under active conditions and their reliability. J Neurosci Methods 2012;205(1):65–71. doi: 10.1016/j.jneumeth.2011.12.012

Ni Z, Gunraj C, Nelson AJ, Yeh IJ, Castillo G, Hoque T, et al. Two phases of interhemispheric inhibition between motor related cortical areas and the primary motor cortex in human. Cereb Cortex 2009;19(7):1654–65. doi: 10.1093/cercor/bhn201

Nielsen CS, Samusyte G, Pugdahl K, Blicher JU, Fuglsang-Frederiksen A, Cengiz B, et al. Test-Retest Reliability of Short-Interval Intracortical Inhibition Assessed by Threshold-Tracking and Automated Conventional Techniques. eNeuro 2021;8(5). doi: 10.1523/ENEURO.0103-21.2021

Nieminen JO, Sinisalo H, Souza VH, Malmi M, Yuryev M, Tervo AE, et al. Multi-locus transcranial magnetic stimulation system for electronically targeted brain stimulation. Brain Stimul 2022;15(1):116–24. doi: 10.1016/j.brs.2021.11.014

O’Leary TJ, Morris MG, Collett J, Howells K. Reliability of single and paired-pulse transcranial magnetic stimulation in the vastus lateralis muscle. Muscle Nerve 2015;52(4):605–15. doi: 10.1002/mus.24584

Oldfield RC. The assessment and analysis of handedness: the Edinburgh inventory. Neuropsychologia 1971;9(1):97–113. doi: 10.1016/0028-3932(71)90067-4

Parmigiani S, Barchiesi G, Cattaneo L. The dorsal premotor cortex exerts a powerful and specific inhibitory effect on the ipsilateral corticofacial system: a dual-coil transcranial magnetic stimulation study. Exp Brain Res 2015;233(11):3253–60. doi: 10.1007/s00221-015-4393-7

Parmigiani S, Zattera B, Barchiesi G, Cattaneo L. Spatial and Temporal Characteristics of Set-Related Inhibitory and Excitatory Inputs from the Dorsal Premotor Cortex to the Ipsilateral Motor Cortex Assessed by Dual-Coil Transcranial Magnetic Stimulation. Brain Topogr 2018;31(5):795–810. doi: 10.1007/s10548-018-0635-x

Pellegrini M, Zoghi M, Jaberzadeh S. The effect of transcranial magnetic stimulation test intensity on the amplitude, variability and reliability of motor evoked potentials. Brain Res 2018;1700:190–8. doi: 10.1016/j.brainres.2018.09.002

Peri E, Ambrosini E, Colombo VM, van de Ruit M, Grey MJ, Monticone M, et al. Intra and inter-session reliability of rapid Transcranial Magnetic Stimulation stimulus-response curves of tibialis anterior muscle in healthy older adults. PLoS One 2017;12(9):e0184828. doi: 10.1016/j.brs.2018.03.002

Picard N, Strick PL. Imaging the premotor areas. Curr Opin Neurobiol 2001;11(6):663–72. doi: 10.1016/s0959-4388(01)00266-5

Portney LG, Watkins MP. Foundations of Clinical Research: Applications to Practice, 3e. New York, NY: McGraw-Hill Education; 2017.

Rodriguez-Nieto G, Seer C, Sidlauskaite J, Vleugels L, Van Roy A, Hardwick R, et al. Inhibition, Shifting and Updating: Inter and intra-domain commonalities and differences from an executive functions activation likelihood estimation meta-analysis. NeuroImage 2022;264:119665. doi: 10.1016/j.neuroimage.2022.119665

Rossini PM, Burke D, Chen R, Cohen LG, Daskalakis Z, Di Iorio R, et al. Non-invasive electrical and magnetic stimulation of the brain, spinal cord, roots and peripheral nerves: Basic principles and procedures for routine clinical and research application. An updated report from an I.F.C.N. Committee. Clin Neurophysiol 2015;126(6):1071–107. doi: 10.1016/j.clinph.2015.02.001

Rothwell JC. Using transcranial magnetic stimulation methods to probe connectivity between motor areas of the brain. Hum Mov Sci 2011;30(5):906–15. doi: 10.1016/j.humov.2010.07.007

Rurak BK, Rodrigues JP, Power BD, Drummond PD, Vallence AM. Test Re-test Reliability of Dual-site TMS Measures of SMA-M1 Connectivity Differs Across Inter-stimulus Intervals in Younger and Older Adults. Neuroscience 2021;472:11–24. doi: 10.1016/j.neuroscience.2021.07.023

Rurak BK, Rodrigues JP, Power BD, Drummond PD, Vallence AM. Reduced Cerebellar Brain Inhibition Measured Using Dual-Site TMS in Older Than in Younger Adults. Cerebellum 2022;21(1):23–38. doi: 10.1007/s12311-021-01267-2

Rushworth MF, Johansen-Berg H, Gobel SM, Devlin JT. The left parietal and premotor cortices: motor attention and selection. NeuroImage 2003;20 Suppl 1:S89–100. doi: 10.1016/j.neuroimage.2003.09.011

Samusyte G, Bostock H, Rothwell J, Koltzenburg M. Short-interval intracortical inhibition: Comparison between conventional and threshold-tracking techniques. Brain Stimul 2018;11(4):806–17. doi: 10.1016/j.brs.2018.03.002

Schambra HM, Ogden RT, Martinez-Hernandez IE, Lin X, Chang YB, Rahman A, et al. The reliability of repeated TMS measures in older adults and in patients with subacute and chronic stroke. Front Cell Neurosci 2015;9:335. doi: 10.3389/fncel.2015.00335

Shibasaki H. Cortical activities associated with voluntary movements and involuntary movements. Clin Neurophysiol 2012;123(2):229–43. doi: 10.1016/j.clinph.2011.07.042

Stewart JC, Dewanjee P, Shariff U, Cramer SC. Dorsal premotor activity and connectivity relate to action selection performance after stroke. Hum Brain Mapp 2016;37(5):1816–30. doi: 10.1002/hbm.23138

Tankisi H, Cengiz B, Howells J, Samusyte G, Koltzenburg M, Bostock H. Short-interval intracortical inhibition as a function of inter-stimulus interval: Three methods compared. Brain Stimul 2021;14(1):22–32. doi: 10.1016/j.brs.2020.11.002

Tracy J, Faro S, Mohammed F, Koenigsberg R, Emperado J. Brain activation change following “overlearned” motor performance. NeuroImage 1998;7(4):S966. doi: 10.1016/s1053-8119(18)31799-3

Turco CV, Pesevski A, McNicholas PD, Beaulieu LD, Nelson AJ. Reliability of transcranial magnetic stimulation measures of afferent inhibition. Brain Res 2019;1723:146394. doi: 10.1016/j.brainres.2019.146394

Vaalto S, Saisanen L, Kononen M, Julkunen P, Hukkanen T, Maatta S, et al. Corticospinal output and cortical excitation-inhibition balance in distal hand muscle representations in nonprimary motor area. Hum Brain Mapp 2011;32(10):1692–703. doi: 10.1002/hbm.21137

Van Hoornweder S, Debeuf R, Verstraelen S, Meesen R, Cuypers K. Unravelling Ipsilateral Interactions Between Left Dorsal Premotor and Primary Motor Cortex: A Proof of Concept Study. Neuroscience 2021;466:36–46. doi: 10.1016/j.neuroscience.2021.04.033

Van Malderen S, Hehl M, Verstraelen S, Swinnen SP, Cuypers K. Dual-site TMS as a tool to probe effective interactions within the motor network: a review. Rev Neurosci 2023;34(2):129–221. doi: 10.1515/revneuro-2022-0020

Vesia M, Barnett-Cowan M, Elahi B, Jegatheeswaran G, Isayama R, Neva JL, et al. Human dorsomedial parieto-motor circuit specifies grasp during the planning of goal-directed hand actions. Cortex 2017;92:175–86. doi: 10.1016/j.cortex.2017.04.007

Volz LJ, Hamada M, Rothwell JC, Grefkes C. What Makes the Muscle Twitch: Motor System Connectivity and TMS-Induced Activity. Cereb Cortex 2015;25(9):2346–53. doi: 10.1093/cercor/bhu032

Wassermann EM. Risk and safety of repetitive transcranial magnetic stimulation: report and suggested guidelines from the International Workshop on the Safety of Repetitive Transcranial Magnetic Stimulation, June 5-7, 1996. Electroencephalogr Clin Neurophysiol 1998;108(1):1–16. doi: 10.1016/s0168-5597(97)00096-8

Wassermann EM. Variation in the response to transcranial magnetic brain stimulation in the general population. Clin Neurophysiol 2002;113(7):1165–71. doi: 10.1016/s1388-2457(02)00144-x

Weir JP. Quantifying test-retest reliability using the intraclass correlation coefficient and the SEM. J Strength Cond Res 2005;19(1):231–40. doi: 10.1519/15184.1

Welniarz Q, Gallea C, Lamy JC, Meneret A, Popa T, Valabregue R, et al. The supplementary motor area modulates interhemispheric interactions during movement preparation. Hum Brain Mapp 2019;40(7):2125–42. doi: 10.1002/hbm.24512

Wessel MJ, Draaisma LR, Morishita T, Hummel FC. The Effects of Stimulator, Waveform, and Current Direction on Intracortical Inhibition and Facilitation: A TMS Comparison Study. Front Neurosci 2019;13:703. doi: 10.3389/fnins.2019.00703

World Medical Association. World Medical Association Declaration of Helsinki: ethical principles for medical research involving human subjects. JAMA 2013;310(20):2191–4. doi: 10.1001/jama.2013.281053

Ziemann U, Rothwell JC, Ridding MC. Interaction between intracortical inhibition and facilitation in human motor cortex. J Physiol 1996;496 (Pt 3)(Pt 3):873–81. doi: 10.1113/jphysiol.1996.sp021734

