## Supplement1-3 for "Test-retest reliability of intrahemispheric dorsal premotor and primary motor cortex dual-site TMS connectivity measures"

^$^Shared first author

^1^Movement Control & Neuroplasticity Research Group, Department of Movement Sciences, Group Biomedical Sciences, KU Leuven, 3001 Heverlee, Belgium

^2^KU Leuven, Leuven Brain Institute (LBI), Leuven, Belgium

^3^Neuroplasticity and Movement Control Research Group, Rehabilitation Research Institute (REVAL), Hasselt University, Diepenbeek, Belgium

***Supplement 1. Quantile-quantile (Q-Q) plots to inspect the normal distribution of standardized residuals as assumed for the relative reliability models.***

Assumption of normally distributed residuals are met in all conditions, except for PMd–M1 stimulation at an ISI of 10ms (Fig. H).

| **A**  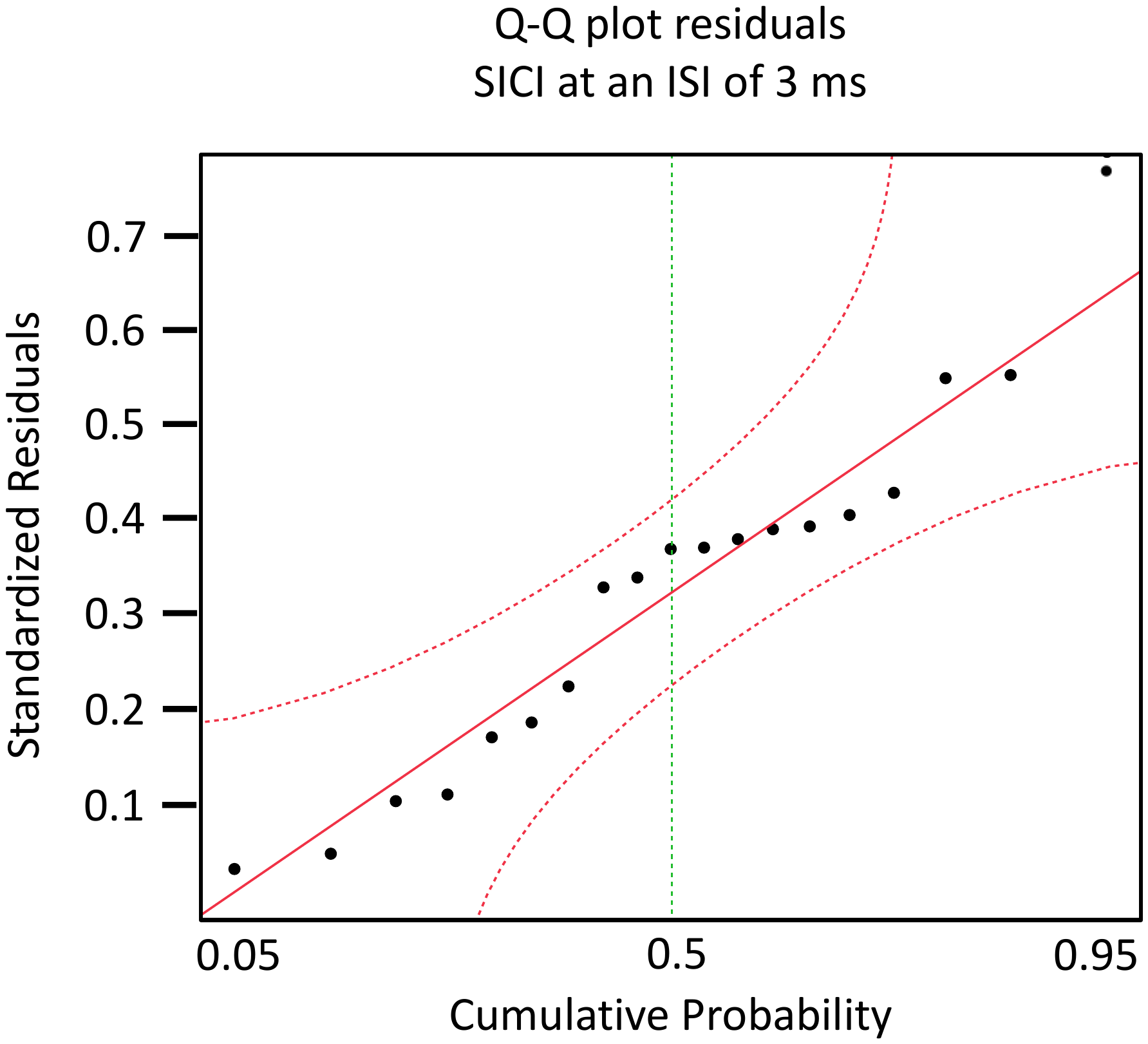 | **B**  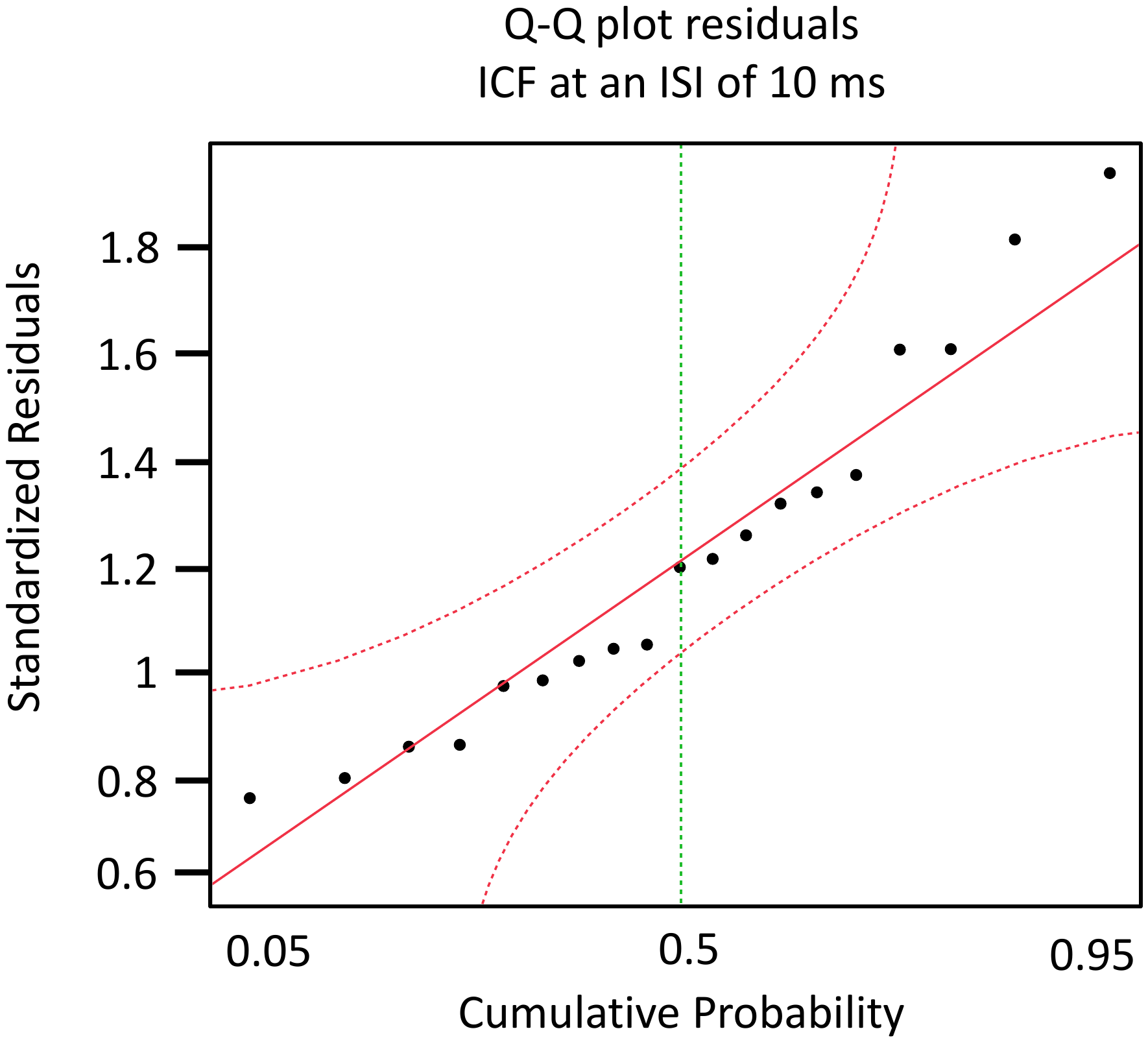 |
| --- | --- |
| **C**  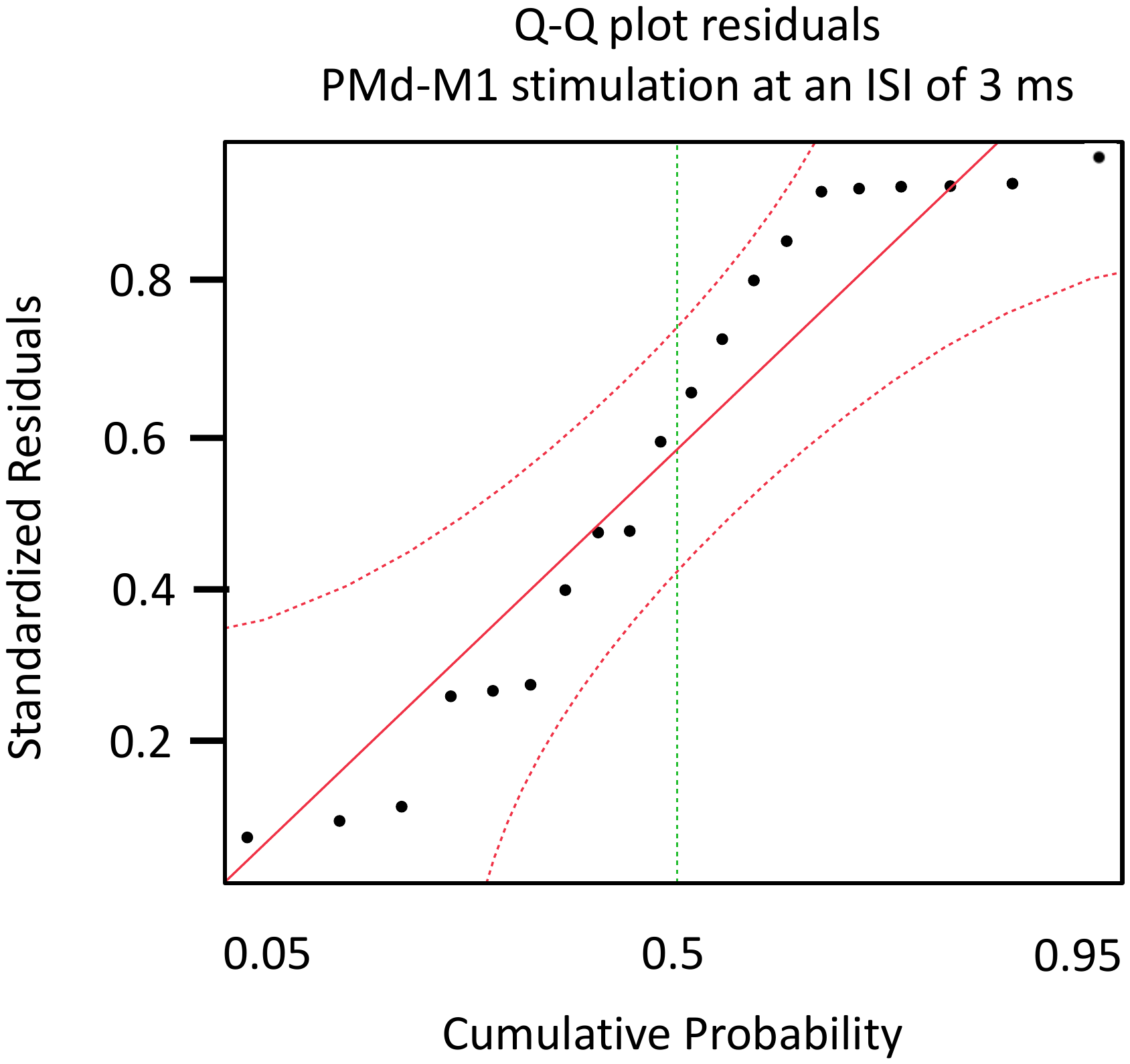 | **D**  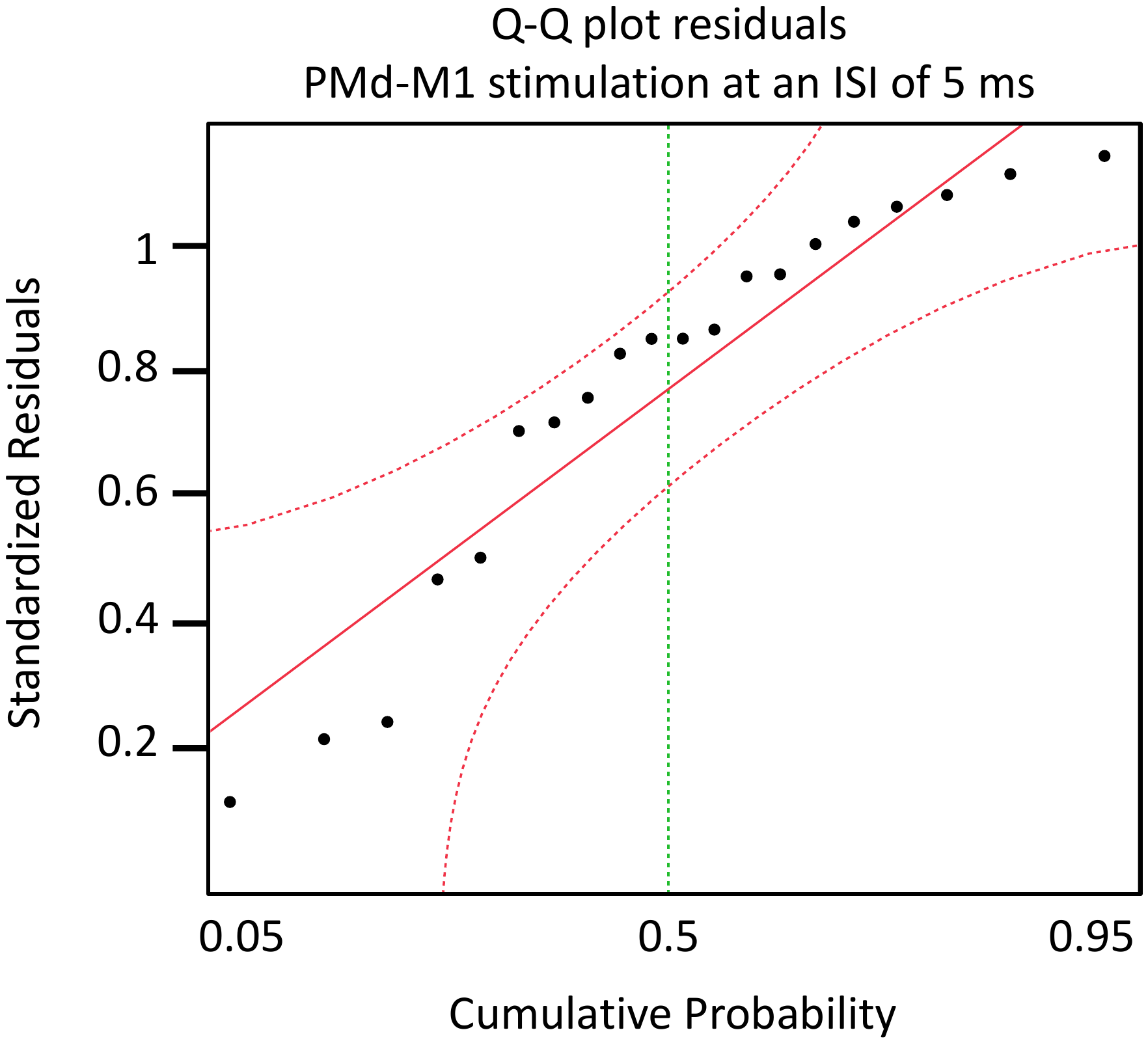 |
| **E**  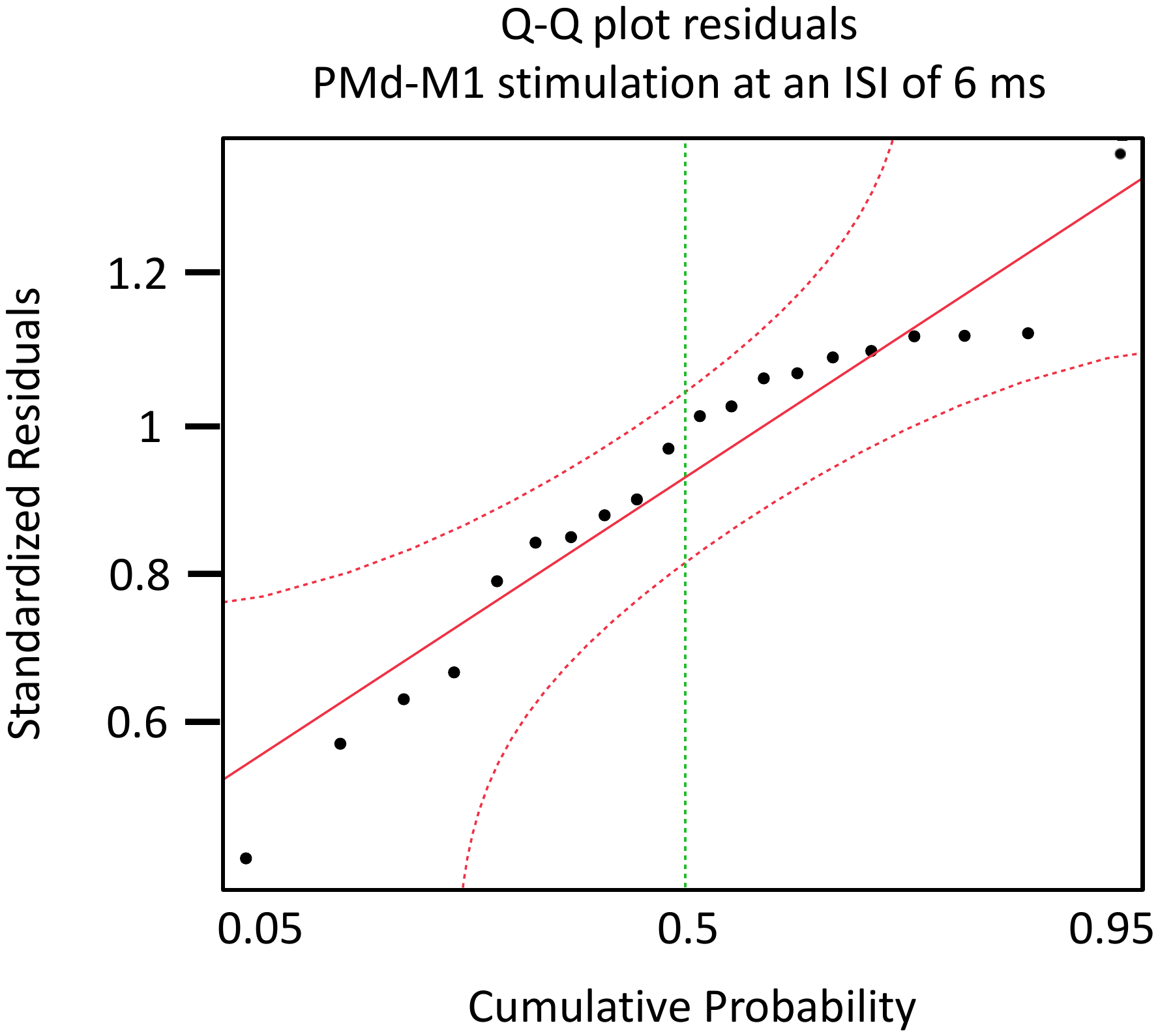 | **F**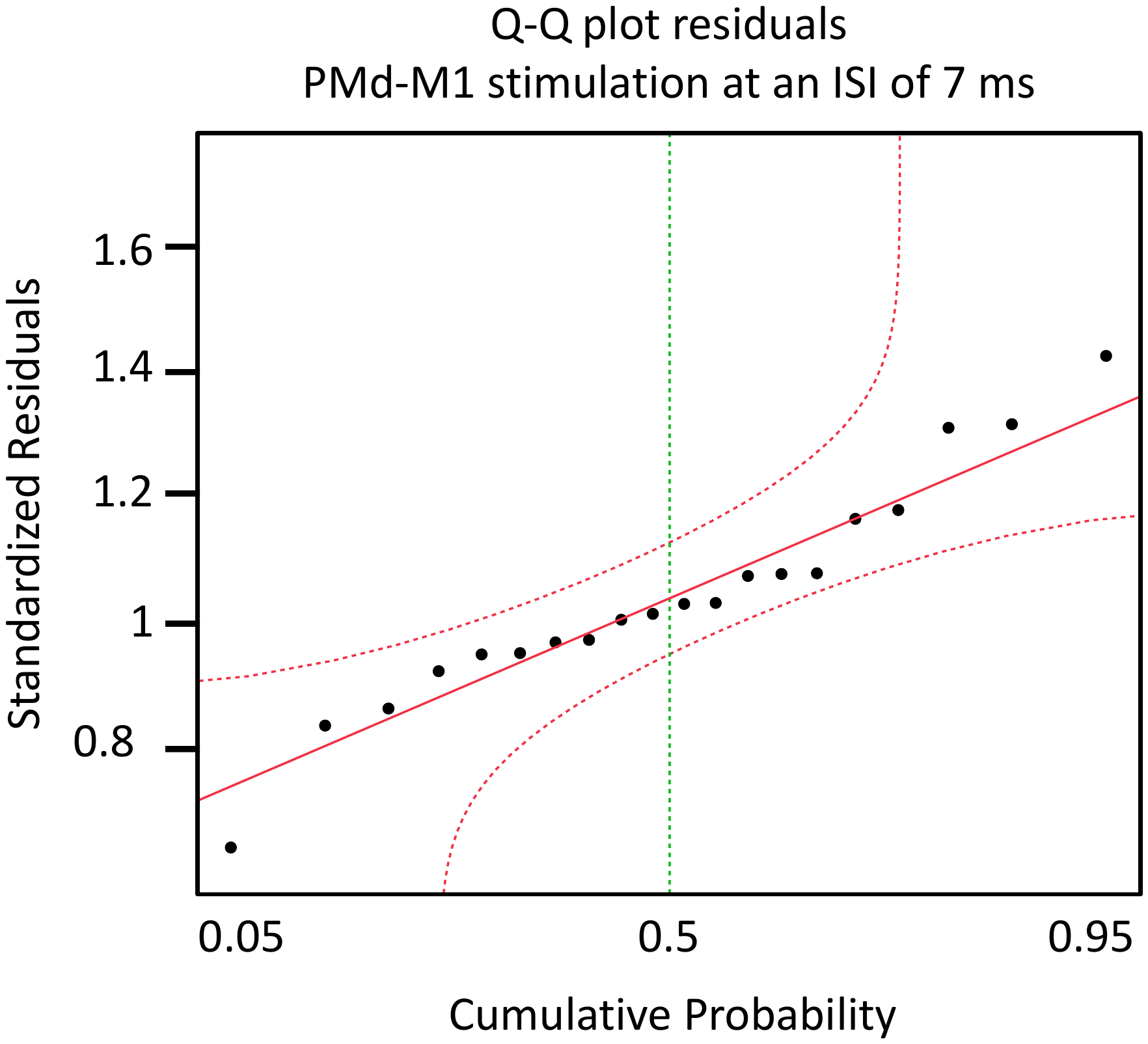 |
| **G**  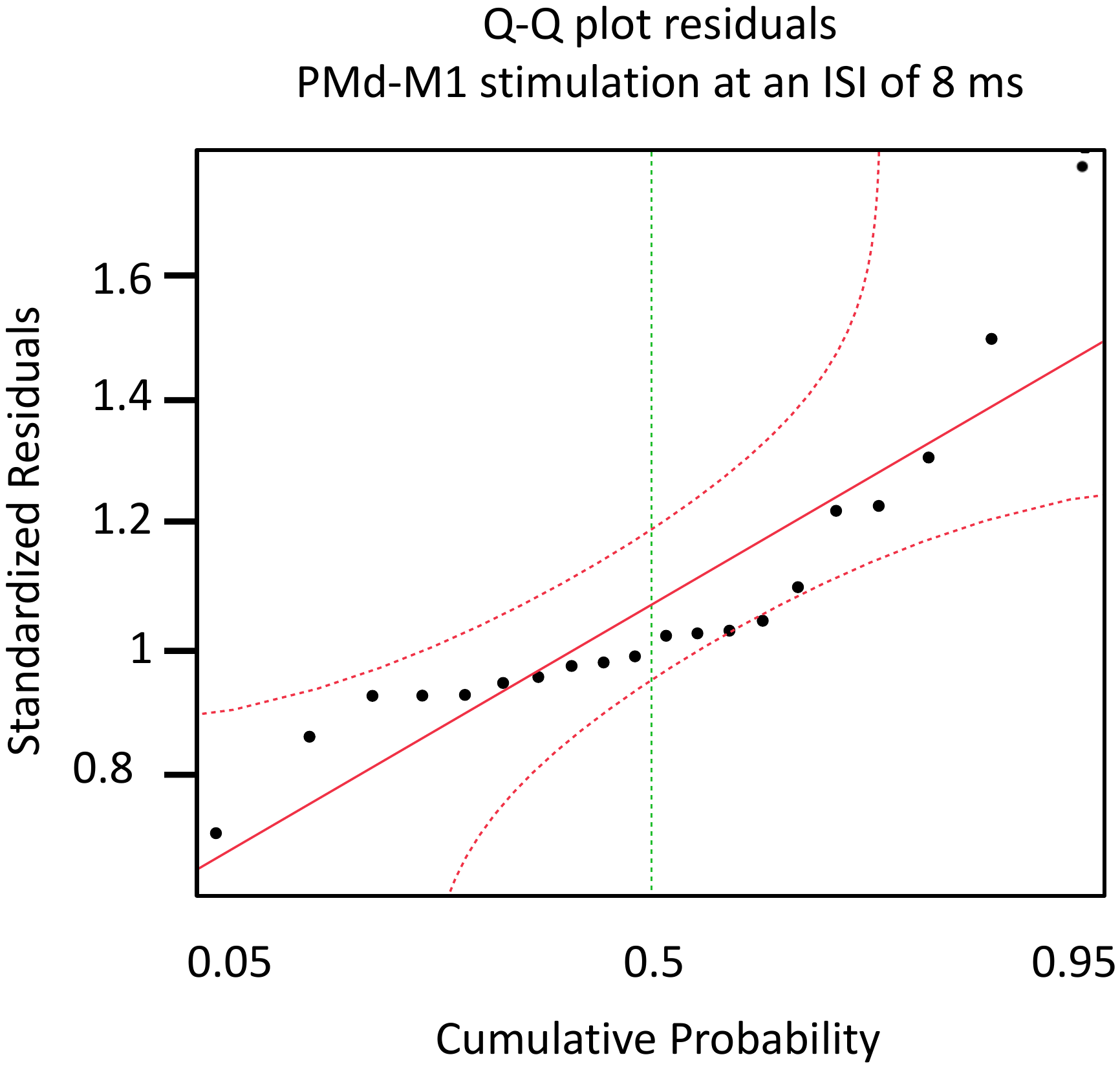 | **H**  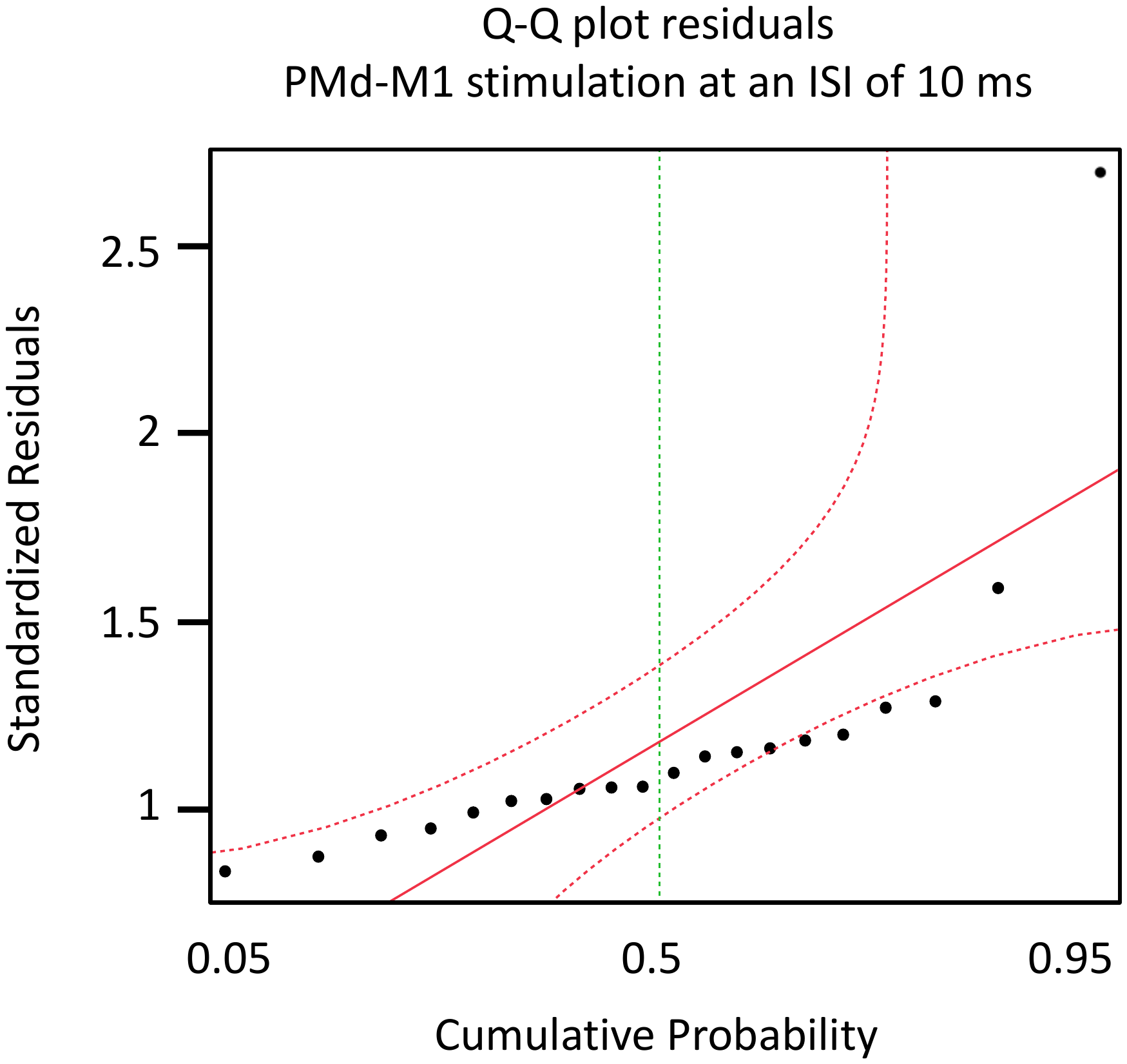 |

Abbreviations: Q-Q plot = quantile-quantile plot; ISI = interstimulus interval; SICI = short intracortical inhibition; ICF = intracortical facilitation; PMd = dorsal pre-motor cortex; M1 = primary motor cortex.

***Supplement 2. Checklist for assessing the methodological quality of studies using transcranial magnetic stimulation to study the motor system (Chipchase et al., 2012).***

The checklist contains 30 items probing the inclusion of correct and complete reporting information related to subjects (8 items), methodology (21 items), and analysis (2 items). Each item could be evaluated as ‘reported’ (i.e., described without further detail) or ‘controlled’ [e.g., excluded, used as a covariate in the analysis, or reported in sufficient (statistical) detail; see Beaulieu et al. (2017)].

| **Were the following participant factors** | **Reported?** | **Controlled?** |
| --- | --- | --- |
| Age of subjects | X | X |
| Gender of subjects | X | **N/A** |
| Handedness of subjects | X | X |
| Subjects prescribed medication | X | X |
| Use of CNS active drugs (e.g. anti-convulsants) | X | X |
| Presence of neurological/psychiatric disorders when studying healthy subjects | X | X |
| Any medical conditions | X | X |
| History of specific repetitive motor activity | - | - |
| **Were the following methodological factors** | | |
| Position and contact of EMG electrodes | X | X |
| Amount of relaxation/contraction of target muscles | X | X |
| Prior motor activity of the muscle to be tested | X | X |
| Level of relaxation of muscles other than those being tested | **N/A** | X |
| Coil type (size and geometry) | X | X |
| Coil orientation | X | X |
| Direction of induced current in the brain | X | X |
| Coil location and stability (with or without a neuronavigation system) | X | X |
| Type of stimulator used (e.g. brand) | X | X |
| Stimulation intensity | X | X |
| Pulse shape (monophasic or biphasic) | X | X |
| Determination of optimal hotspot | X | X |
| The time between MEP trials | X | X |
| Time between days of testing | X | X |
| Subject attention (level of arousal) during testing | X | X |
| Method for determining threshold (active/resting) | X | X |
| Number of MEP measures made | X | X |
| Paired pulse only: Intensity of test pulse | X | X |
| Paired pulse only: Intensity of conditioning pulse | X | X |
| Paired pulse only: Inter-stimulus interval | X | X |
| **Were the following analytical factors** | | |
| Method for determining MEP size during analysis | X | X |
| Size of unconditioned MEP | X | X |

$$\boldsymbol{Total score=}\frac{\boldsymbol{Reported+Controlled}}{\boldsymbol{58}}\boldsymbol{=96.5\%}$$

***Supplement 3. Boxplots visualizing the similarity across TMS measures between measurement days.***

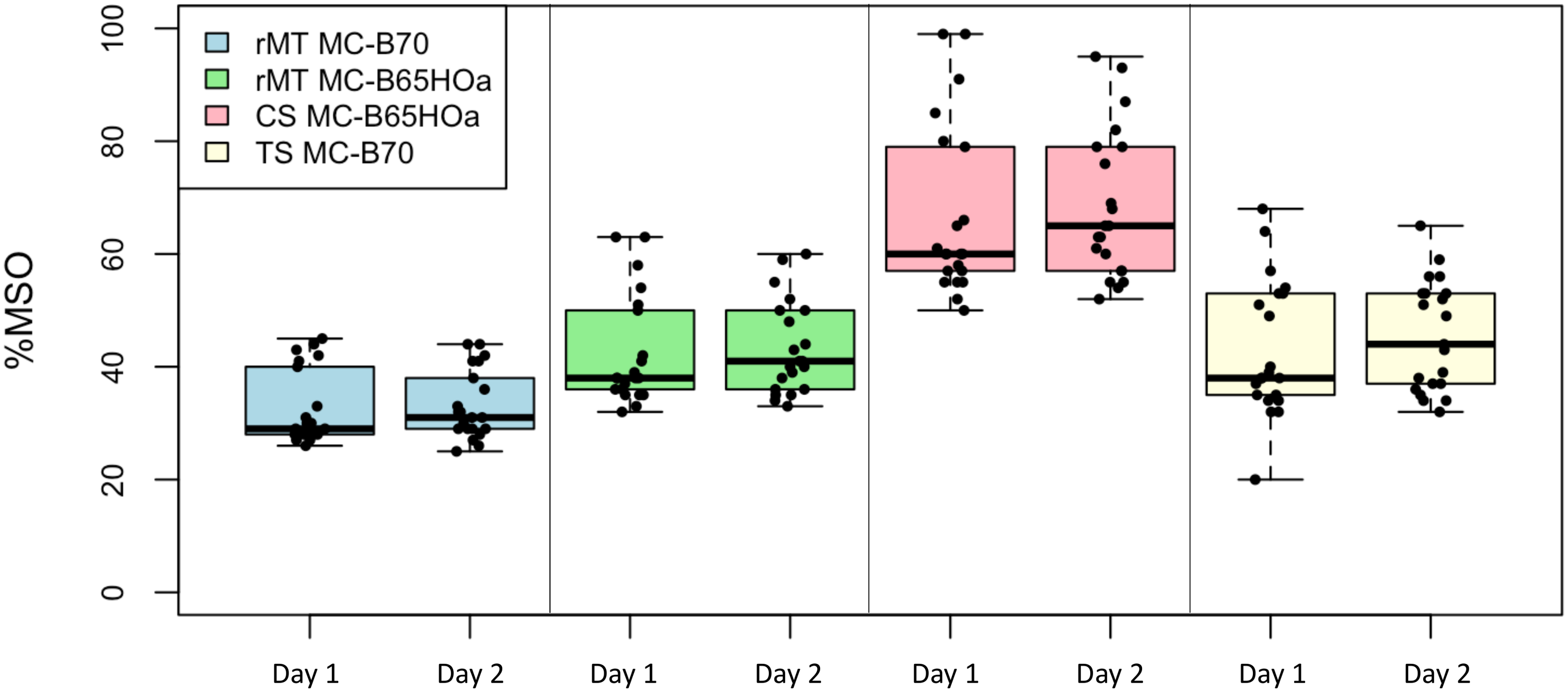

Abbreviations: MT = resting motor threshold; CS = conditioning stimulus; TS = test stimulus; % MSO = percentage maximal stimulator output.

**References:**

Beaulieu, L. D., Flamand, V. H., Masse-Alarie, H., & Schneider, C. (2017). Reliability and minimal detectable change of transcranial magnetic stimulation outcomes in healthy adults: A systematic review. *Brain Stimul*, *10*(2), 196-213. <https://doi.org/10.1016/j.brs.2016.12.008>

Chipchase, L., Schabrun, S., Cohen, L., Hodges, P., Ridding, M., Rothwell, J., Taylor, J., & Ziemann, U. (2012). A checklist for assessing the methodological quality of studies using transcranial magnetic stimulation to study the motor system: an international consensus study. *Clin Neurophysiol*, *123*(9), 1698-1704. <https://doi.org/10.1016/j.clinph.2012.05.003>
